# Optimal Surveillance Strategies for Barrett’s Esophagus Based on Segment Length and Dysplasia Grade: A Cost-Effectiveness Study

**DOI:** 10.64898/2026.02.17.26346313

**Authors:** Akiko Kowada

## Abstract

The risk of esophageal adenocarcinoma (EAC) in Barrett’s esophagus (BE) varies substantially by segment length and dysplasia grade. This study evaluated the cost-effectiveness and health impacts of dysplasia-stratified EAC surveillance strategies for the Japanese BE population. A state-transition model was developed comparing endoscopy, sponge test, breath test, and miRNA test with no surveillance from a healthcare payer perspective over a lifetime. Non-invasive strategies were assessed as primary surveillance tools, with positive results triggering confirmatory endoscopy, and a scenario analysis evaluated AI-assisted endoscopy. Five BE populations of 50-year-old individuals were modeled: ultra-short segment BE (USSBE), short-segment BE (SSBE), long-segment nondysplastic BE (LSBE-NDBE), LSBE with low-grade dysplasia (LSBE-LGD), and LSBE with high-grade dysplasia (LSBE-HGD). Each modality was evaluated at surveillance intervals of 1, 2, 3, 4, 5, or 10 years. Primary outcomes included net monetary benefits, costs, quality-adjusted life-years, incremental cost-effectiveness ratios, and EAC deaths, with sensitivity analyses assessing parameter uncertainty. Surveillance was not cost-effective for USSBE, SSBE, or LSBE-NDBE. For LSBE-LGD, annual endoscopy was most cost-effective, averting 83 EAC deaths per 10,000 individuals, while for LSBE-HGD, annual breath testing was most cost-effective, averting 295 deaths. These findings support dysplasia-specific surveillance in LSBE with implications for global surveillance practice.

## 1. Introduction

Barrett’s esophagus (BE) is the principal precursor to esophageal adenocarcinoma (EAC), a malignancy with rising incidence and poor prognosis when diagnosed at advanced stages. Globally, EAC accounted for approximately 85,700 new cases in 2020, representing about 14% of all esophageal cancers, with the highest incidence observed in high-income regions such as North America, Western and Northern Europe, and Australia [1]. EAC began increasing sharply in Europe and the United States in the 1960s, marking the beginning of a rapid epidemiological transition [2]. In contrast, EAC remains relatively uncommon across Asia; however, the prevalence of BE has increased in several East Asian countries over the past two decades. In Japan, although squamous cell carcinoma remains predominant, the incidence of EAC has shown a gradual upward trend, suggesting that Japan may be entering a phase similar to that experienced by Western countries several decades earlier [3].

In Western countries, major gastroenterological societies —including the American College of Gastroenterology (ACG) [4], the American Society for Gastrointestinal Endoscopy (ASGE) [5], and the British Society of Gastroenterology (BSG) [6]—recommend structured endoscopic surveillance for patients with BE. Surveillance intervals are primarily determined by dysplasia grade, with segment length used to further refine intervals in nondysplastic BE (NDBE). Alongside these guideline-based strategies, several emerging technologies aim to enhance early detection of dysplasia and EAC. These innovations serve two complementary roles: improving the diagnostic performance of endoscopy, exemplified by artificial Intelligence (AI)-assisted systems capable of real-time identification of subtle neoplastic changes [7]; and reducing the burden of endoscopic examinations through minimally invasive pre-endoscopic triage tools such as sponge-based cell-collection devices (e.g., Cytosponge) [8], breath-analysis platforms that detect volatile organic compounds [9], and circulating biomarker assays including miRNA-based tests [10].

In Japan, however, a standardized national surveillance framework for BE has not been established. Although BE is frequently detected in Japan, surveillance practices vary widely among institutions, and most cases represent ultrashort-segment BE (USSBE) [3]. This high detection rate reflects Japan’s sensitive and precise endoscopic diagnostic practices, supported by a broader endoscopic definition that classifies any visible columnar epithelium as BE. Recent epidemiological analyses suggest that the incidence of EAC in Japan is projected to increase in the coming decades [3]. Japan has long maintained a nationwide endoscopic screening program for gastric cancer, providing regular opportunities for incidental detection of esophageal neoplasia among individuals over 50 years of age. As gastric cancer incidence continues to decline due to decreasing *Helicobacter pylori* infection rates and widespread eradication therapy [11], accompanied by a marked increase in reflux esophagitis, the current screening system is expected to be simplified. This may reduce opportunities for incidental EAC detection, even if gastric cancer surveillance is maintained after *H. pylori* eradication. Consequently, establishing a dedicated and evidence-based surveillance strategy for BE has become increasingly important.

Given the rising prevalence of BE, the anticipated increase in EAC incidence, and the absence of a unified surveillance strategy in Japan, the cost-effectiveness of stratified EAC surveillance approaches warrants rigorous evaluation. Therefore, the aim of this study is to evaluate the cost-effectiveness of optimal stratified EAC surveillance strategies for the BE population in Japan, with surveillance intervals defined by segment length and dysplasia grade.

## 2. Methods

### 2.1 Model Overview

We developed a state-transition model to evaluate the cost-effectiveness of stratified surveillance strategies for EAC among individuals with BE in Japan. Five BE subpopulations were modeled independently based on segment length and dysplasia grade: ultra-short segment BE (USSBE), short-segment BE (SSBE), long-segment BE (LSBE) without dysplasia (LSBE-NDBE), LSBE with low-grade dysplasia (LSBE-LGD), and LSBE with high-grade dysplasia (LSBE-HGD) (Figure 1). Each subgroup was simulated separately to reflect its distinct annual incidence of EAC [12,13].

**Figure 1.**
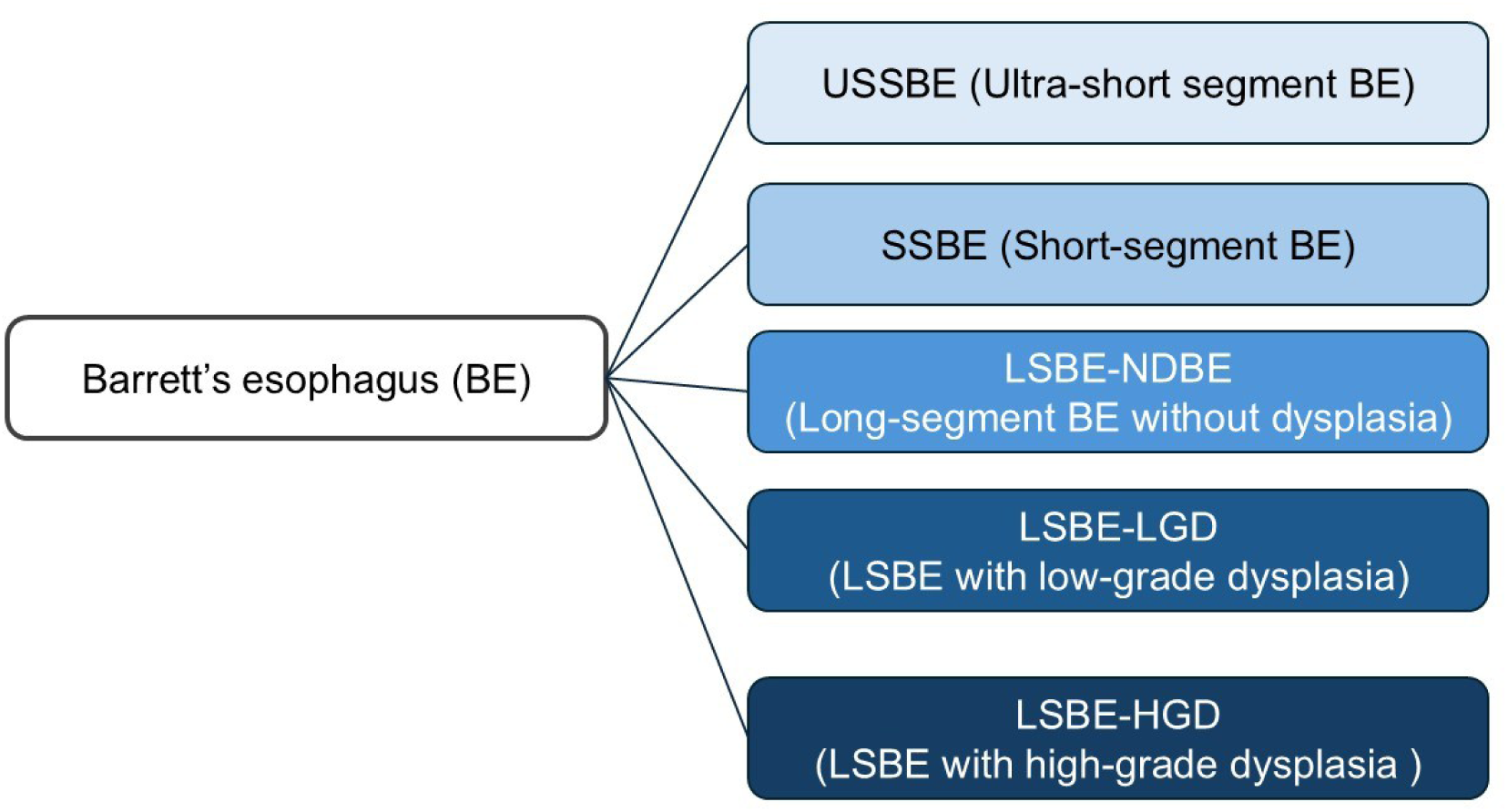
Classification of Barrett’s esophagus subtypes. Visual representation of Barrett’s esophagus (BE) subtypes, categorized by segment length and dysplasia status. The diagram distinguishes ultrashort-segment BE (USSBE), short-segment BE (SSBE), long-segment BE (LSBE) with nondysplastic BE (LSBE-NDBE), low-grade dysplasia (LGD), and high-grade dysplasia (HGD).

The model followed a hypothetical cohort of 50-year-old individuals with BE from a healthcare payer perspective over a lifetime. Age 50 was selected because it approximates the median age at BE diagnosis and allows evaluation of lifetime surveillance benefits. Surveillance strategies included conventional endoscopy, sponge test, breath test, and miRNA test, each evaluated at intervals of 1, 2, 3, 4, 5, or 10 years. Non-invasive strategies were modeled as primary surveillance tools followed by confirmatory endoscopy, and a no-surveillance strategy served as the reference comparator. A one-year cycle length with half-cycle correction was applied, and both costs and quality-adjusted life-years (QALYs) were discounted at 3% annually.

### 2.2 Model Structure

Five independent Markov models were constructed, one for each BE subgroup, to reflect subgroup-specific EAC incidence without assuming unobserved natural-history transitions. Individuals entered the model in their corresponding BE health state (NDBE, LGD, or HGD), underwent surveillance according to the assigned strategy, and remained in that state unless they developed EAC or died.

No transitions between BE states were assumed because the model relied solely on observed EAC incidence and mortality rather than hypothetical natural-history progression pathways. No preclinical disease state or interval cancer construct was included; all cancers were assumed to occur and be diagnosed within the same annual cycle. Mortality incorporated stage-specific EAC survival [14] and background mortality from Japanese life tables [15]. Primary outcomes included net monetary benefits (NMBs), costs, QALYs, incremental cost-effectiveness ratios (ICERs), and EAC deaths. A schematic representation of health states and allowable transitions is shown in Figure 2.

**Figure 2.**
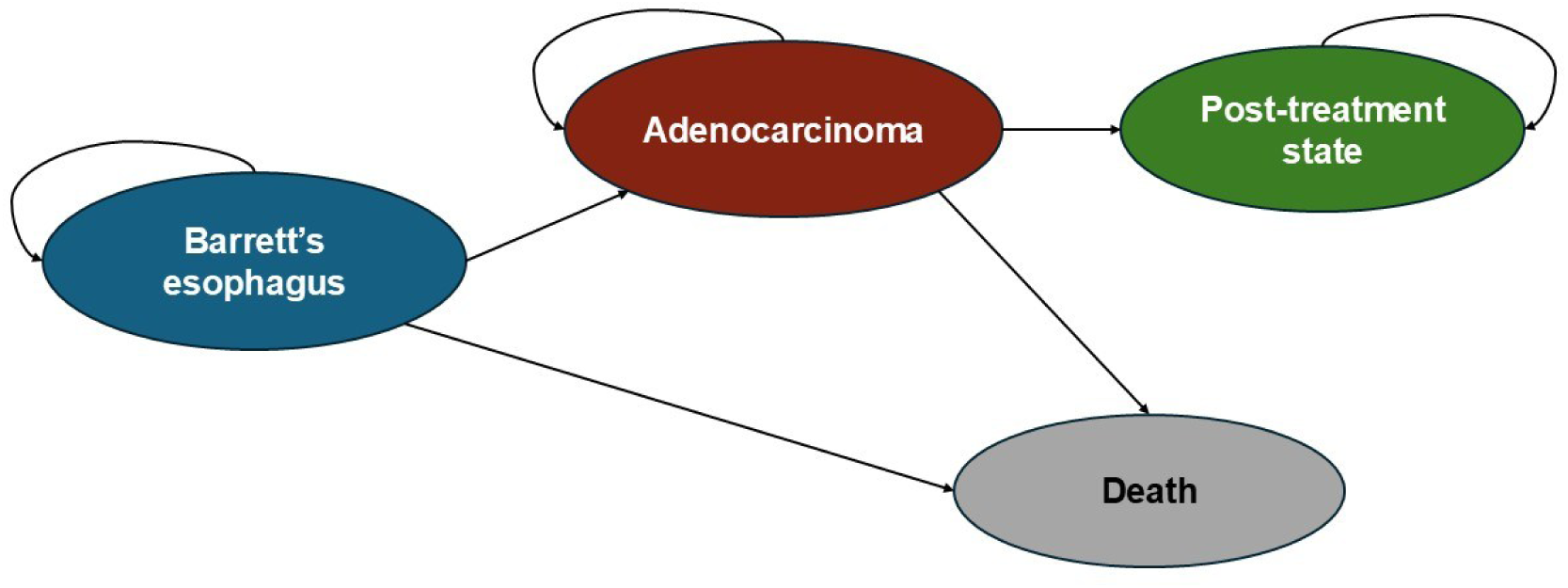
State-transition diagram for Barrett’s esophagus progression. Markov model structure illustrating disease progression and outcomes for patients with Barrett’s esophagus. Health states include Barrett’s esophagus, esophageal adenocarcinoma, post-treatment state, and death. Arrows indicate possible transitions between states, including progression to esophageal adenocarcinoma, treatment, and death.

### 2.3 Surveillance Strategies

Four surveillance modalities were evaluated: conventional endoscopy, sponge test, breath test, and miRNA test. Each modality was assessed at surveillance intervals of 1, 2, 3, 4, 5, or 10 years, with no surveillance as the reference. AI-assisted endoscopy was evaluated in scenario analyses as an enhanced form of conventional endoscopy.

Conventional endoscopy represented the current standard of care in Japan. Sponge test, breath test, and miRNA test were modeled as non-invasive primary surveillance tools. A positive non-endoscopic test triggered confirmatory endoscopy and subsequent management according to stage-specific treatment pathways in the Japanese Guidelines for the Diagnosis and Treatment of Esophageal Cancer [16]. Individuals with negative results continued routine surveillance at the assigned interval.

### 2.4 Diagnostic Performance

Sensitivity and specificity for conventional endoscopy were obtained from meta-analyses and large observational cohorts [17]. Sensitivity and specificity for sponge test, breath test, miRNA test, and AI-assisted endoscopy were extracted from published studies [8,9,10,18].

False-negative results were modeled as cancers diagnosed clinically within the same cycle, with stage distribution based on clinically detected EAC. Adherence rates for conventional endoscopy, AI-assisted endoscopy, and breath test were obtained from published sources [19], whereas adherence for sponge test and miRNA test was assumed. All adherence parameters were varied widely (10–90%) in sensitivity analyses.

### 2.5 Model Inputs

Model inputs included subgroup-specific annual EAC incidence [12,13], diagnostic performance parameters, costs, health-state utilities, and mortality rates. Annual EAC incidence was assigned separately for each BE subgroup (USSBE, SSBE, LSBE-NDBE, LSBE-LGD, LSBE-HGD). Sensitivity, specificity, and adherence were assigned independently for each surveillance modality [8,9,10,17,18]. Costs reflected testing and cancer treatment expenditures [20]. Health-state utilities were derived from published literature [21,22]. Stage-specific EAC survival was based on Japanese data [14], and background mortality was derived from Japanese life tables [15]. Costs and QALYs were discounted at 3% annually [23].

#### 2.5.1 Costs

Costs were derived from the Japanese national fee schedule [20] and converted to U.S. dollars using the 2024 OECD purchasing power parity exchange rate (95.1 yen per US$1) [24]. Costs for sponge test, breath test, AI-assisted endoscopy, and miRNA test were not available in Japan and were therefore treated as assumptions, with ranges explored in sensitivity analyses.

#### 2.5.2 Health-State Utilities

Utilities were assigned to mutually exclusive health states representing the clinical course of BE and EAC, including NDBE, LGD, HGD, four stage-specific EAC states (I–IV), and three post-treatment states [21,22]. Death was modeled as an absorbing state. Utility values were derived from published literature and discounted at 3% [23].

### 2.6 Base-Case Analysis

The base-case analysis estimated costs, QALYs, ICERs, and NMBs. Strategies were ordered by increasing QALYs, and extended dominance was applied when appropriate. A willingness-to-pay threshold of US$50,000 per QALY gained was used to determine cost-effectiveness [25].

### 2.7 Markov Cohort Analyses

Using a Markov cohort model, we estimated, for each surveillance strategy, the cumulative 10-year and lifetime numbers of EAC deaths averted and the cumulative 10-year and lifetime additional costs per EAC death averted, compared with no surveillance. Analyses were conducted separately for hypothetical cohorts of 10,000 LSBE-LGD patients and 10,000 LSBE-HGD patients aged 50 years.

For each cohort, EAC deaths averted per 10,000 patients were calculated by multiplying the difference in the cumulative probability of EAC death between each surveillance strategy and no surveillance by 10,000. Additional costs per EAC death averted were calculated by multiplying the difference in total discounted costs between each surveillance strategy and no surveillance by 10,000 and dividing this value by the number of EAC deaths averted in the corresponding 10,000-patient cohort.

### 2.8 Scenario Analyses

Scenario analyses evaluated AI-assisted endoscopy by substituting its diagnostic performance parameters for those of conventional endoscopy within each model. This approach allowed us to explore how improvements in sensitivity and specificity attributable to AI would influence overall cost-effectiveness across the surveillance strategies.

### 2.9 Sensitivity Analyses

One-way sensitivity analyses varied key clinical and economic parameters across plausible ranges, including test sensitivity and specificity, surveillance and treatment costs, health-state utilities, and subgroup-specific EAC incidence. We also varied the starting age of surveillance from 40 to 80 years to evaluate how age at initiation influenced cost-effectiveness outcomes. Deterministic results were summarized using tornado diagrams.

A probabilistic sensitivity analysis was conducted using 10,000 Monte Carlo simulations. Parameter uncertainty was represented using beta distributions for test performance, subgroup-specific EAC incidence, and utilities, and gamma distributions for costs. For each simulation, lifetime costs and QALYs were recalculated, and cost-effectiveness acceptability curves were generated at a willingness-to-pay (WTP) threshold of US$50,000 per QALY gained.

All analyses were conducted using TreeAge Pro 2026 (TreeAge Software, Williamstown, MA). This economic evaluation followed the CHEERS 2022 reporting guidelines [26].

### 2.10 Model Validation

Model validation included face, internal, and external validation. Face validation assessed whether the model structure, assumptions, and parameter inputs were consistent with established frameworks for BE and EAC modeling. Internal validation evaluated whether simulated transitions, incidence patterns, and long-term outcomes behaved as expected across all five BE subgroups. External validation compared model-generated estimates of EAC incidence, stage distribution, and survival with published epidemiologic data. These checks supported the credibility of the model for evaluating stratified surveillance strategies.

## 3. Results

### 3.1 Base-case results

Base-case outcomes differed markedly across BE subgroups, driven primarily by variation in annual EAC incidence.

#### USSBE

No surveillance was the preferred strategy, yielding the highest NMB (US$869,600) (Table S1). Annual endoscopy increased costs substantially (US$3,942 vs US$28) with only negligible QALY gains, lowering NMB to US$865,697. Annual breath test and annual miRNA test produced even lower NMBs.

#### SSBE

No surveillance again produced the highest NMB (US$950,019) (Table S2). Annual endoscopy increased costs (US$4,117 vs US$225) with minimal QALY improvement, reducing NMB to US$946,232. Across both short-segment groups, surveillance strategies increased costs with only marginal benefits and were not cost-effective at a WTP of US$50,000 per QALY.

#### LSBE-NDBE

No surveillance remained the preferred strategy (NMB US$938,821) (Table S3, Figure 3A). Annual endoscopy raised costs (US$5,484 vs US$1,770) and modestly improved QALYs, but resulted in a lower NMB (US$935,939). Annual breath test and annual miRNA test yielded even lower NMBs. None of the surveillance strategies were cost-effective.

**Figure 3.**
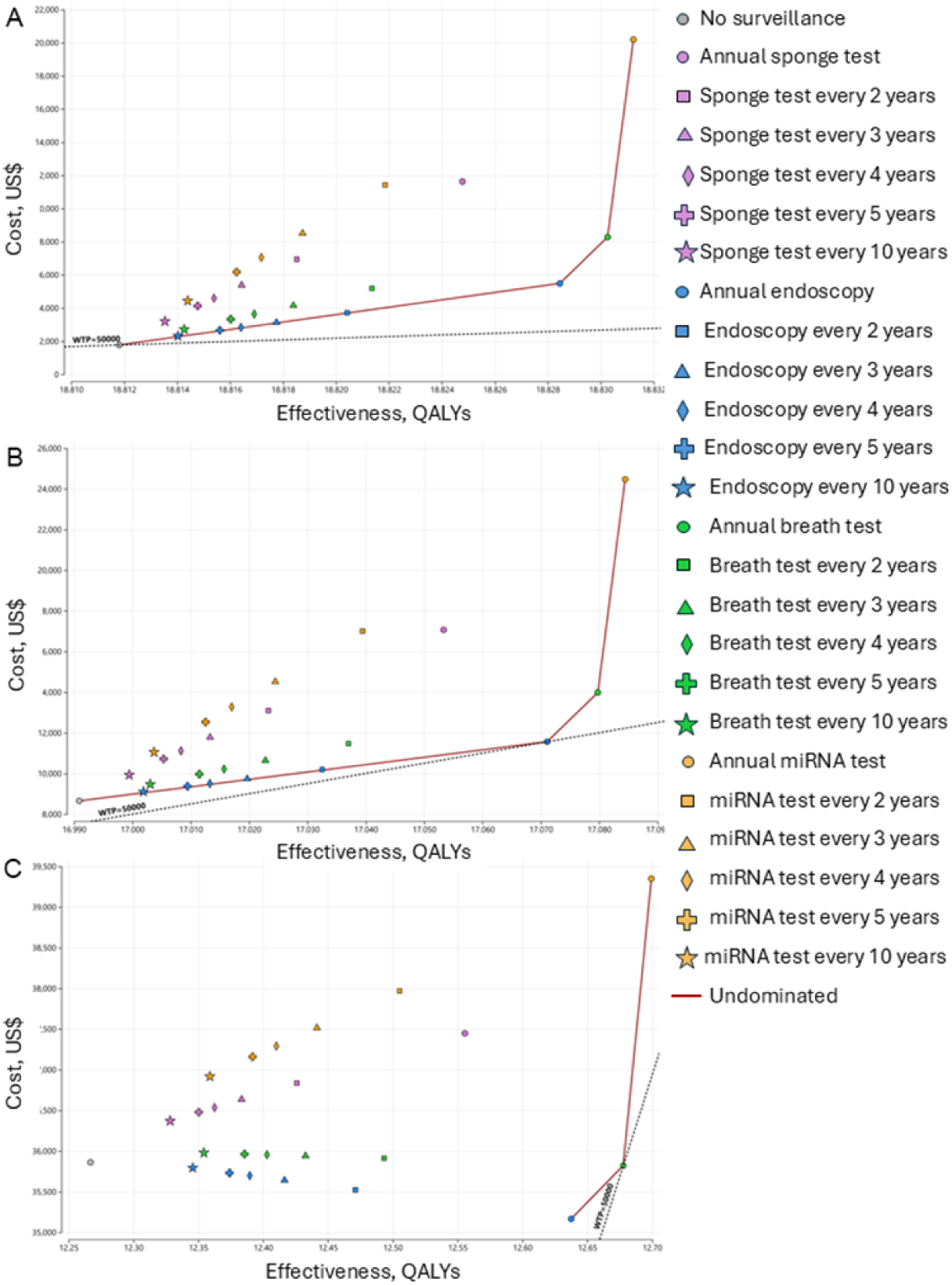
Base-case results for LSBE subgroups. A. LSBE with nondysplastic Barrett’s esophagus (LSBE-NDBE) B. LSBE with low-grade dysplasia (LSBE-LGD) C. LSBE with high-grade dysplasia (LSBE-HGD) Base-case analysis comparing screening strategies in patients with LSBE, stratified by dysplasia status. Each panel presents incremental cost and effectiveness for evaluated screening strategies. The dashed line indicates the willingness-to-pay threshold of US$50,000 per QALY. Strategies on the cost-effectiveness frontier are considered undominated. LSBE, long-segment Barrett’s esophagus.

#### LSBE-LGD

Annual endoscopy was the most cost-effective strategy, producing the highest NMB (US$841,993) (Table S4, Figure 3B). Although no surveillance had lower costs, it also produced fewer QALYs, resulting in a slightly lower NMB. Annual breath test and annual miRNA test generated marginally higher QALYs but substantially lower NMBs, indicating that their additional benefits did not justify their higher costs.

#### LSBE-HGD

Annual breath test was the preferred strategy, with the highest NMB (US$598,066) and an ICER of US$16,318 per QALY gained (Table S5, Figure 3C). Annual miRNA test produced the highest QALYs but was not cost-effective due to its high ICER and lower NMB.

#### Overall

Surveillance was not cost-effective in low-risk subgroups (USSBE, SSBE, LSBE-NDBE). In contrast, annual endoscopy for LSBE-LGD and annual breath test for LSBE-HGD offered the greatest value in higher-risk populations.

### 3.2 EAC deaths averted and additional costs per death averted

EAC deaths averted and additional costs per EAC death averted were evaluated for 10,000 LSBE-LGD and 10,000 LSBE-HGD patients over 10 years and a lifetime.

In LSBE-LGD, annual endoscopy prevented 24 EAC deaths within 10 years and 83 EAC deaths over a lifetime (Table 2, Figure S6), with lifetime additional costs of US$349,308 per EAC death averted (Table 3, Figure S8). Annual breath test and annual miRNA test prevented slightly more EAC deaths (92 and 97 lifetime deaths, respectively) but required substantially higher additional costs per EAC death averted.

**Table 1.** Baseline estimates for selected variables.

| Variable | Baseline value | One-way sensitivity analysis range | Distribution | Reference |
| --- | --- | --- | --- | --- |
| **Probabilities** |  |  |  |  |
| Annual EAC incidence in USSBE | 0.000032 | 0.0000066-0.00013 | Beta | 12 |
| Annual EAC incidence in SSBE | 0.00026 | 0.00011-0.00054 | Beta | 12 |
| Annual EAC incidence in LSBE-NDBE | 0.0021 | 0.0012-0.0029 | Beta | 13 |
| Annual EAC incidence in LSBE-LGD | 0.0116 | 0.0057-0.0175 | Beta | 13 |
| Annual EAC incidence in LSBE-HGD | 0.1416 | 0.1243-0.1589 | Beta | 13 |
| Adherence rate (%) |  |  |  |  |
| Endoscopy | 78 | 10-100 | Beta | 19 |
| AI-assisted endoscopy | 78 | 10-100 | Beta | 19 |
| Sponge test | 75 | 10-100 | Beta | 19 |
| Breath test | 95 | 10-100 | Beta | Assumption |
| miRNA test | 95 | 10-100 | Beta | Assumption |
| Sensitivity (%) |  |  |  |  |
| Endoscopy | 75 | 60-100 | Beta | 17 |
| AI-assisted endoscopy | 93 | 88-100 | Beta | 18 |
| Sponge test | 81 | 76-85 | Beta | 8 |
| Breath test | 91 | 84-95 | Beta | 9 |
| miRNA test | 95.8 | 89.8-98.4 | Beta | 10 |
| Specificity (%) |  |  |  |  |
| Endoscopy | 70 | 55-95 | Beta | 17 |
| AI-assisted endoscopy | 78 | 53-94 | Beta | 18 |
| Sponge test | 89 | 82-93 | Beta | 8 |
| Breath test | 74 | 69-79 | Beta | 9 |
| miRNA test | 95.2 | 87.1-98.4 | Beta | 10 |
| **Costs (US\$) US\$1 = ¥95.1** | | | | |
| Endoscopy | 234 | 117-468 | Gamma | 20 |
| AI-assisted endoscopy | 256 | 128-512 | Gamma | Assumption |
| Sponge test | 420 | 210-840 | Gamma | Assumption |
| Breath test | 60 | 30-120 | Gamma | Assumption |
| miRNA test | 700 | 350-1,400 | Gamma | Assumption |
| EAC stage I treatment | 7,361 | 3,681-14,722 | Gamma | 20 |
| EAC stage II treatment | 28,391 | 14,196-56,782 | Gamma | 20 |
| EAC stage III treatment | 10,515 | 5,258-21,030 | Gamma | 20 |
| EAC stage IV treatment | 21,030 | 10,515-42,060 | Gamma | 20 |
| **Health state utilities** |  |  |  |  |
| NDBE | 0.93 | 0.88-0.96 | Beta | 21,22 |
| LGD | 0.88 | 0.82-0.92 | Beta |  |
| HGD | 0.82 | 0.75-0.88 | Beta |  |
| EAC stage I | 0.78 | 0.72-0.84 | Beta |  |
| EAC stage II | 0.72 | 0.65-0.78 | Beta |  |
| EAC stage III | 0.62 | 0.55-0.70 | Beta |  |
| EAC stage IV | 0.50 | 0.40-0.60 | Beta |  |
| Post-treatment state NDBE | 0.93 | 0.88-0.96 | Beta |  |
| Post-treatment state LGD | 0.88 | 0.82-0.92 | Beta |  |
| Post-treatment state HGD | 0.82 | 0.75-0.88 | Beta |  |
| Death | 0 | N/A | - |  |

AI, artificial intelligence; EAC, esophageal adenocarcinoma; NDBE, non-dysplastic Barrett's esophagus; LGD, low-grade dysplasia; HGD, high-grade dysplasia; USSBE, ultrashort-segment Barrett's esophagus; SSBE, short-segment Barrett's esophagus; LSBE, long-segment Barrett's esophagus; miRNA, microRNA; N/A, not applicable.

**Table 2.**
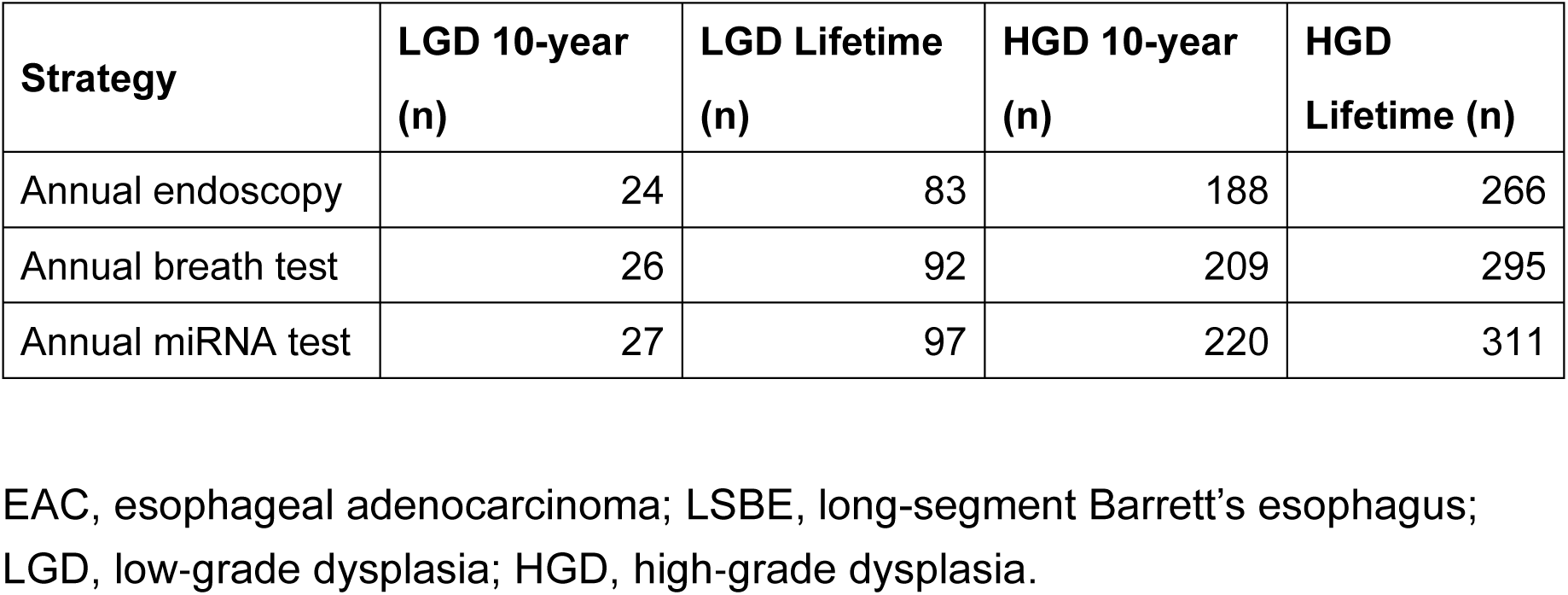
EAC deaths averted per 10,000 LSBE-LGD and 10,000 LSBE-HGD patients over 10 years and over a lifetime.

| Strategy | LGD 10-year (n) | LGD Lifetime (n) | HGD 10-year (n) | HGD Lifetime (n) |
| --- | --- | --- | --- | --- |
| Annual endoscopy | 24 | 83 | 188 | 266 |
| Annual breath test | 26 | 92 | 209 | 295 |
| Annual miRNA test | 27 | 97 | 220 | 311 |

EAC, esophageal adenocarcinoma; LSBE, long-segment Barrett’s esophagus; LGD, low-grade dysplasia; HGD, high-grade dysplasia.

**Table 3.**
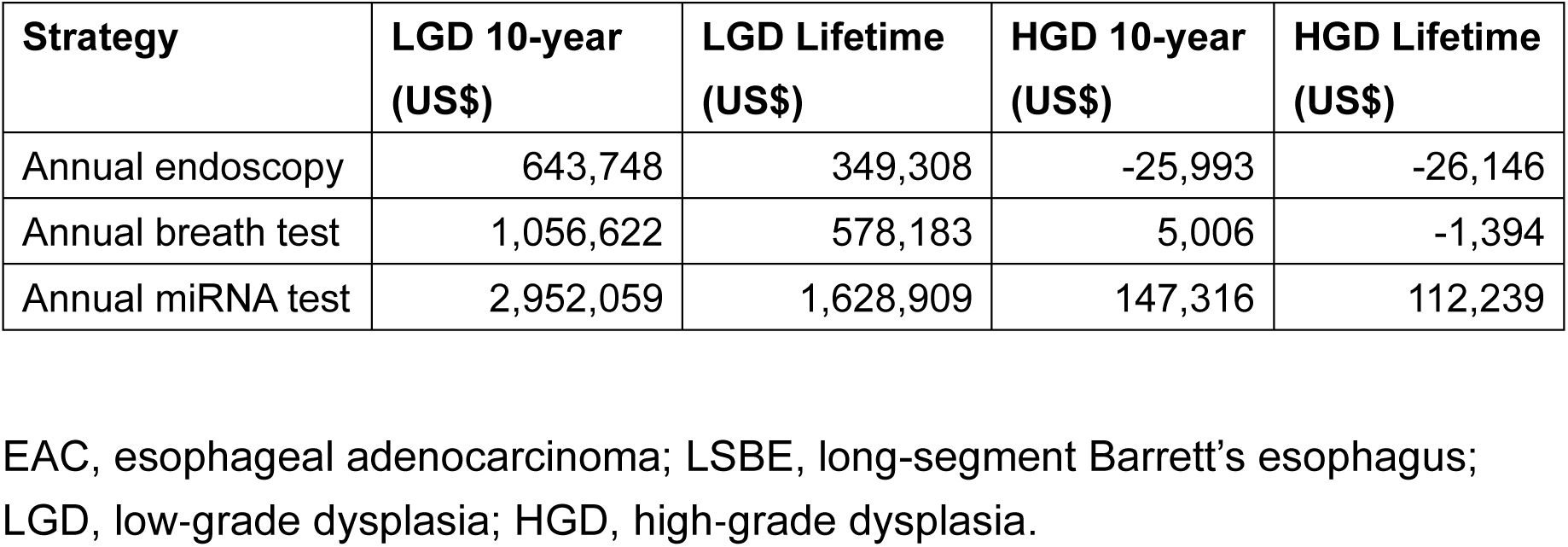
Additional costs per EAC death averted among 10,000 LSBE-LGD and 10,000 LSBE-HGD patients over 10 years and over a lifetime.

In LSBE-HGD, annual breath test prevented 209 EAC deaths within 10 years and 295 deaths over a lifetime (Table 2, Figure S7), costing US$5,006 over 10 years but ultimately saving US$1,394 over a lifetime per EAC death averted (Table 3, Figure S9). Although annual miRNA test prevented the most deaths (311 over a lifetime), it did so at substantially higher additional costs per EAC death averted. These findings indicate that the absolute mortality benefit of surveillance rises sharply with baseline EAC incidence, with annual breath test offering the most favorable balance of EAC death reduction and cost in HGD.

### 3.3 Scenario analyses

When AI-assisted endoscopy was evaluated as an alternative to conventional endoscopy, no surveillance remained cost-effective in USSBE, SSBE, and LSBE-NDBE at a WTP threshold of US$50,000 per QALY (Table S5–S8).

In LSBE-LGD, annual AI-assisted endoscopy yielded the highest NMB (US$842,747), whereas in LSBE-HGD, annual breath test following AI-assisted endoscopy produced the highest NMB (US$603,334) (Table S9 and Table S10). These effects were driven by the higher sensitivity and specificity of AI-assisted endoscopy, which increased early detection while reducing false positives in higher-risk subgroups.

### 3.4 Sensitivity analyses

One-way sensitivity analyses indicated that cost-effectiveness was primarily driven by subgroup-specific EAC incidence and test adherence.

In LSBE-LGD, annual endoscopy became cost-effective relative to no surveillance when annual EAC incidence exceeded 0.009 (Figure S1).

In LSBE-HGD, annual breath test was favored over annual endoscopy when endoscopy adherence exceeded 83.5% and breath-test adherence remained below 88.8% (Figure S2). Annual miRNA test became more cost-effective than annual breath test when breath-test adherence fell below 83.7% (Figure S3).

Cost-effectiveness acceptability curves showed a 79% probability that no surveillance is cost-effective in LSBE-NDBE (Figure 4A), a 36% probability that annual endoscopy is cost-effective in LSBE-LGD (Figure 4B and Figure S4), and a 29% probability that annual breath test is cost-effective in LSBE-HGD (Figure 4C and Figure S5) at a WTP threshold of US$50,000 per QALY gained.

**Figure 4.**
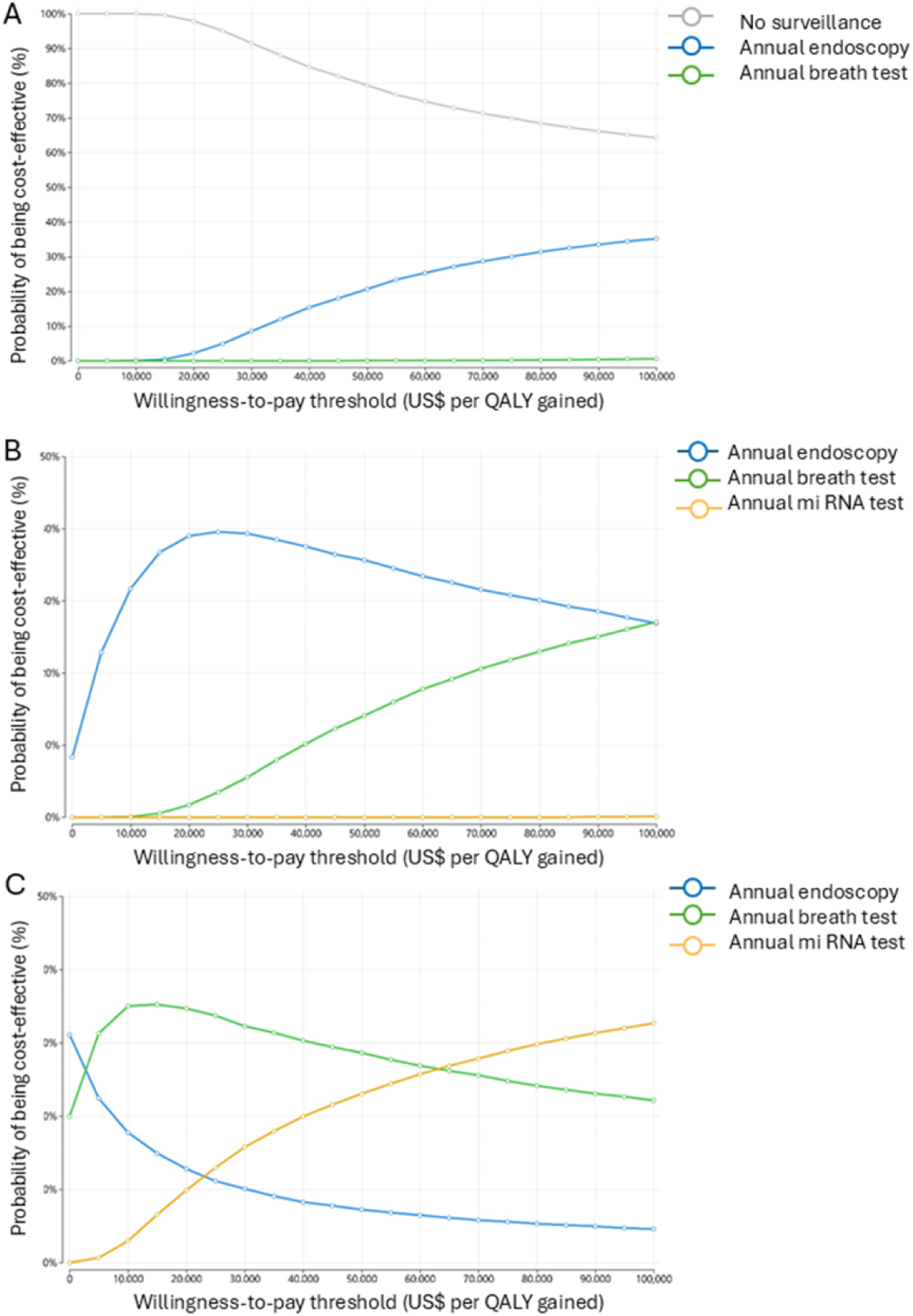
Cost-effectiveness acceptability curves for LSBE subgroups. A. LSBE with nondysplastic Barrett’s esophagus (LSBE-NDBE) B. LSBE with low-grade dysplasia (LSBE-LGD) C. LSBE with high-grade dysplasia (LSBE-HGD) Curves show the probability that each screening strategy is cost-effective across a range of willingness-to-pay thresholds, based on probabilistic sensitivity analyses. Colors are consistent across all panels to represent the same screening strategies. LSBE, long-segment Barrett’s esophagus.

#### Robustness of Findings Across Subgroups

Findings on which subgroups benefit from surveillance were generally robust across sensitivity analyses, with low-risk subgroups consistently favoring no surveillance. Although the overall decision to perform surveillance remained stable, the relative ranking of surveillance strategies within higher-risk subgroups (LGD or HGD) shifted depending on subgroup-specific EAC incidence and test adherence.

## 4. Discussion

### 4.1 Principal Findings

This study demonstrates that stratified surveillance strategies for BE, defined by segment length and dysplasia grade, provide the most cost-effective approach for the Japanese population. By incorporating Japan-specific incidence data for USSBE and SSBE and applying Western subgroup-specific incidence estimates for LSBE (NDBE, LGD, and HGD) derived from meta-analyses [12], we developed a transparent incidence-based framework across five clinically distinct subgroups. Using this framework, we identified that annual endoscopy is the optimal strategy for patients with LGD, whereas annual breath test is the most cost-effective strategy for those with HGD.

Across subgroups, differences in EAC deaths averted closely mirrored differences in baseline EAC incidence, indicating that incidence is the primary determinant of the preferred surveillance strategy. As incidence increased, the optimal strategy shifted in a clear stepwise pattern: surveillance offered limited value in USSBE, SSBE, and LSBE-NDBE; annual endoscopy became optimal in LSBE-LGD; and annual breath testing became optimal in LSBE-HGD. This pattern reflects the strong dependence of surveillance benefit on underlying cancer risk and the diagnostic performance of available tests.

Because AI-assisted endoscopy improves sensitivity and specificity from 75% and 70% to 93% and 78%, respectively, it directly replaces standard endoscopy in the incidence range where endoscopic surveillance was previously optimal. This shift illustrates how improvements in test accuracy can move the incidence thresholds that define the preferred strategy, further reinforcing the transparency and adaptability of an incidence-based modeling approach.

To our knowledge, this study provides the first fully integrated, risk-stratified cost-effectiveness evaluation of EAC surveillance in BE worldwide. It also incorporates both conventional and AI-assisted endoscopic modalities, as well as emerging non-endoscopic approaches. Importantly, because LSBE incidence inputs are derived from Western literature, the model is not limited to Japan; substituting region-specific cost parameters would allow direct application to Western healthcare settings.

Japan provides a unique context for evaluating surveillance strategies because it has one of the world’s highest proportions of ultrashort-segment BE [3] and a lower baseline incidence of EAC compared with Western countries. Historically, early detection of EAC in Japan has relied on incidental findings during gastric cancer screening, routine health checkups, and other clinical encounters. As opportunities for incident detection are expected to decline [3], risk-stratified surveillance becomes increasingly important for maintaining early-stage diagnosis while avoiding unnecessary procedures in low-risk groups. Emerging technologies such as breath test and AI-assisted endoscopy may further enhance surveillance efficiency, particularly in intermediate- and high-risk groups. Overall, this study provides a policy-relevant foundation for developing surveillance recommendations tailored to Japan while offering a generalizable framework applicable to Western populations.

### 4.2 Comparison with Previous Studies

Two cost-effectiveness analyses have previously evaluated BE surveillance. Kastelein et al. demonstrated that surveillance with endoscopic mucosal resection and radiofrequency ablation for HGD or early EAC, and oesophagectomy for advanced EAC, is cost-effective every 5 years for NDBE and every 3 years for LGD, based on a Dutch healthcare perspective and a willingness-to-pay threshold of €35,000 per QALY [27]. Vissapragada et al. further identified risk-stratified surveillance strategies as the most cost-effective, showing that biennial surveillance for long-segment BE (>2 cm) and 12-month surveillance for LGD provided the best value, thereby confirming the utility of dysplasia-based risk stratification in Australia [28]. These cost-effectiveness models simulated the natural history from BE to EAC and required numerous structural assumptions, including progression rates, dwell times, and transition probabilities, that vary widely across studies and contribute to inconsistent estimates of surveillance benefit, particularly in NDBE. Our findings additionally identified annual breath test for LSBE-HGD and annual endoscopy for LSBE-LGD as the most cost-effective strategies, and showed that AI-assisted endoscopy can serve as a cost-effective alternative to conventional endoscopy. Our study also employs an incidence-based framework anchored to empirically observed annual EAC incidence, avoiding assumptions about preclinical disease dynamics and providing a more transparent representation of cancer risk across clinically distinct subgroups.

Evaluating the cost-effectiveness of surveillance for USSBE and SSBE is essential because these segments represent the majority of BE cases in Japan [3]. This study is the first to demonstrate limited cost-effectiveness of surveillance for NDBE—including USSBE and SSBE—when empirical incidence is applied without additional structural assumptions. Our model also incorporates Western dysplasia-specific incidence estimates for LSBE [13], reflecting the limited availability of Japanese data and anticipating future increases in LGD and HGD, thereby supporting applicability to Western populations as well. Furthermore, our evaluation of emerging technologies, such as breath test and AI-assisted endoscopy, extends previous work by demonstrating their potential value in Japan and in higher-incidence settings [9,18]. Non-endoscopic examinations may further alleviate the surveillance burden for individuals with BE, particularly in settings where frequent endoscopy is difficult to sustain.

### 4.3 Strengths and Limitations

This study has several methodological strengths. First, it employs an incidence-based framework that does not rely on unobservable progression states or assumptions about transitions between nondysplastic BE, dysplasia, and preclinical cancer. By grounding the model in empirically observed annual EAC incidence for each subgroup, the analysis avoids the structural uncertainty inherent in traditional multistate models and provides a transparent representation of cancer risk. Second, evaluating five clinically distinct BE subgroups independently allows surveillance strategies to be compared within risk strata, clarifying why surveillance yields limited benefit in low-risk groups and substantial benefit in high-risk groups. Third, the model incorporates modality-specific test performance and applies consistent probability distributions across parameters, enhancing transparency and reducing reliance on unverifiable assumptions. Finally, using a fixed cohort of 10,000 individuals per subgroup ensures comparability across analyses, allowing surveillance strategies to be evaluated independently of uncertain subgroup size estimates.

This study also has limitations. First, although Japan has published EAC incidence estimates for USSBE, SSBE, and LSBE overall [12], dysplasia-stratified incidence estimates are not available. Further research will be needed to establish reliable incidence estimates for LSBE subgroups, including separate estimates for NDBE, LGD, and HGD. Second, reliable data on the population size of each BE subgroup in Japan are lacking [3], which prevented estimation of national-level impacts and necessitated the use of a fixed cohort size for subgroup analyses. Third, several evaluated modalities, such as sponge test, breath test, miRNA test, and AI-assisted endoscopy, are not yet reimbursed under the Japanese national insurance system, requiring cost assumptions that may differ from future reimbursement prices. Fourth, test performance parameters were derived from heterogeneous Western studies [8,9,10,17,18], and real-world performance in Japan may differ. However, this limitation was addressed by conducting sensitivity analyses using wide parameter ranges. Finally, patient burden associated with undergoing surveillance tests was not incorporated into the model, although it is an important consideration, particularly for non-invasive modalities.

### 4.4 Clinical and Policy Implications

The findings of this study have important implications for clinical practice and health policy. First, the clear stratification of surveillance benefit across Barrett’s esophagus subgroups supports a shift toward risk-based surveillance rather than uniform intervals. The limited value of surveillance in USSBE, SSBE, and LSBE-NDBE suggests that routine endoscopic surveillance may not be justified for these low-risk groups in Japan. Conversely, the strong cost-effectiveness observed in LSBE-LGD and LSBE-HGD reinforces the need for timely and intensive surveillance in higher-risk patients, consistent with their substantially elevated incidence of EAC.

Second, the results highlight the potential role of emerging non-endoscopic technologies, particularly breath test, which demonstrated favorable cost-effectiveness in high-risk groups. These modalities may reduce the burden of repeated endoscopy, improve patient acceptability, and expand access to surveillance in settings with limited endoscopic capacity. AI-assisted endoscopy may further enhance diagnostic accuracy and efficiency as evidence accumulates [7]. AI-assisted endoscopy, which surpasses the diagnostic performance of conventional endoscopy and offers greater cost-effectiveness, is likely to replace standard endoscopy in the incidence range where it is currently preferred, further improving the efficiency of surveillance programs.

Third, future integration of individualized risk-prediction tools could further enhance precision. Although segment length and dysplasia grade remain the strongest determinants of cancer risk, additional factors, such as age, sex, obesity, and GERD severity [29,30], could refine risk stratification and guide personalized surveillance intensity.

Fourth, the incidence-based and hybrid structure of the model, which uses Japan-specific incidence for USSBE and SSBE and Western subgroup-specific incidence for LSBE, ensures that the framework is not restricted to Japan and can be directly applied to other regions, including Western countries, by substituting local cost parameters. This generalizability enhances the relevance of the findings beyond Japan and provides a foundation for international comparisons of surveillance strategies.

Fifth, structured risk-stratified surveillance will become increasingly important for maintaining early-stage EAC detection while avoiding unnecessary procedures in low-risk groups, particularly as EAC incidence is expected to rise in Japan. Policymakers may consider incorporating these findings into future national guidelines to optimize resource allocation and improve patient outcomes.

Finally, integrating BE/EAC surveillance into Japan’s upper gastrointestinal screening system and post–*H. pylori* eradication surveillance could allow these pathways to be restructured into a more comprehensive upper gastrointestinal screening framework. Such integration may improve efficiency in lower-risk populations while maintaining appropriate monitoring for higher-risk patients.

## 5. Conclusions

This study demonstrates that the value of surveillance for Barrett’s esophagus varies substantially across subgroups defined by segment length and dysplasia grade. Annual endoscopy is cost-effective for LGD, whereas annual breath test is cost-effective for HGD. In contrast, surveillance offers limited value for NDBE. These findings support prioritizing surveillance resources for higher-risk groups while avoiding unnecessary procedures in low-risk populations.

By grounding the model in empirically observed cancer incidence and avoiding assumptions about unobservable progression states, this study provides a transparent and reproducible framework for evaluating surveillance strategies. Its incidence-based Markov structure makes the framework broadly generalizable and applicable beyond Japan, including to Western healthcare systems.

Emerging technologies, including AI-assisted endoscopy, sponge test, breath test, and miRNA test, may further enhance the cost-effectiveness of surveillance, particularly in intermediate- and high-risk groups. The development of non-endoscopic procedures is also expected to reduce patient burden and expand access to surveillance.

As EAC incidence is projected to rise in Japan, these findings offer timely, population-specific evidence to guide future surveillance recommendations. Adoption of dysplasia-stratified, incidence-aligned strategies may help optimize resource allocation while maximizing clinical benefit in Japan and could serve as a model for other regions with similarly low but increasing EAC incidence.

## List of Abbreviations

AI: Artificial intelligence
BE: Barrett’s esophagus
EAC: Esophageal adenocarcinoma
HGD: High-grade dysplasia
ICER: Incremental cost-effectiveness ratio
LGD: Low-grade dysplasia
LSBE: Long-segment Barrett’s esophagus
miRNA: MicroRNA
NDBE: Nondysplastic Barrett’s esophagus
NMB: Net monetary benefit
QALY: Quality-adjusted life-year
SSBE: Short-segment Barrett’s esophagus
USSBE: Ultrashort-segment Barrett’s esophagus

## Acknowledgments

None.

## Data availability

All data used in this study are publicly available aggregated national statistics, and no individual-level data were used.

## Funding

This research received no specific grant from any funding agency in the public, commercial, or not-for-profit sectors.

## Competing interests

The author declares no competing interests.

## Ethics statement

This study used publicly available, aggregated national statistics and did not involve human subjects. Therefore, ethical approval was not required.

## Author contributions

The author conceptualized the study, developed the model, conducted the analysis, validated the model, interpreted the results, drafted the manuscript, and revised the final version.

## Notes

### Competing Interest Statement

The authors have declared no competing interest.

### Funding Statement

This study did not receive any funding.

### Author Declarations

National cancer statistics published by the Ministry of Health, Labour and Welfare (Japan) Published epidemiological parameters from peer-reviewed literature Publicly available demographic data from national statistical agencies

### Summary of Updates

The title was refined for clarity. Several sentences in the main text were revised to improve readability. Supplementary materials were updated and reorganized for accuracy.

## Reference

1. Morgan, E. et al. The global landscape of esophageal squamous cell carcinoma and esophageal adenocarcinoma incidence and mortality in 2020 and projections to 2040: New estimates from GLOBOCAN 2020. Gastroenterology 163, 649–658.e2 (2022). 10.1053/j.gastro.2022.05.054

2. Blot, W. J., Devesa, S. S., Kneller, R. W. & Fraumeni, J. F. Rising incidence of adenocarcinoma of the esophagus and gastric cardia. JAMA 265, 1287–1289 (1991).

3. Iijima, K. Epidemiology of Barrett’s neoplasia in Japan. Digestion 107, 5–14 (2026). 10.1159/000548362

4. Shaheen, N. J. et al. ACG clinical guideline: Diagnosis and management of Barrett’s esophagus. Am. J. Gastroenterol. 111, 30–51 (2016). 10.1038/ajg.2015.322

5. Qumseya, B. et al. ASGE guideline on screening and surveillance of Barrett’s esophagus. Gastrointest. Endosc. 90, 335–359.e2 (2019). 10.1016/j.gie.2019.05.012

6. Fitzgerald, R. C. et al. British Society of Gastroenterology guidelines on the diagnosis and management of Barrett’s oesophagus. Gut 63, 7–42 (2014). 10.1136/gutjnl-2013-305372

7. Hashimoto, R. et al. Artificial intelligence using convolutional neural networks for real-time detection of early esophageal neoplasia in Barrett’s esophagus. Gastrointest. Endosc. 91, 1264–1271.e1 (2020). 10.1016/j.gie.2019.12.049

8. Kakar, M. T. et al. Diagnostic test accuracy of Cytosponge–Trefoil Factor 3 for Barrett esophagus: A systematic review and meta-analysis. JGH Open 9, e70248 (2025). 10.1002/jgh3.70248

9. Peters, Y. et al. Detection of Barrett’s oesophagus through exhaled breath using an electronic nose device. Gut 69, 1169–1172 (2020). 10.1136/gutjnl-2019-320273

10. Miyoshi, J. et al. Liquid biopsy to identify Barrett’s oesophagus, dysplasia and oesophageal adenocarcinoma: The EMERALD multicentre study. Gut 74, 169–181 (2025). 10.1136/gutjnl-2024-333364

11. Kowada, A. & Asaka, M. Economic and health impacts of introducing Helicobacter pylori eradication strategy into national gastric cancer policy in Japan: A cost-effectiveness analysis. Helicobacter 26, e12837 (2021). 10.1111/hel.12837

12. Fukuda, S. et al. Cancer risk by length of Barrett’s esophagus in Japanese population: A nationwide multicenter retrospective cohort study. J. Gastroenterol. 59, 887–895 (2024). 10.1007/s00535-024-02139-2

13. Tan, J. et al. Incidence rates of Barrett’s esophagus and esophageal adenocarcinoma: A systematic review and meta-analysis. iGIE 3, 92–103 (2024). 10.1016/j.igie.2024.01.001

14. Cancer Information Service, National Cancer Center Japan. Cancer registry and statistics. https://ganjoho.jp/reg_stat/statistics/stat/cancer/5_stomach.html (Accessed 14 Feb 2026).

15. Ministry of Health, Labour and Welfare Japan. Vital statistics. https://www.mhlw.go.jp/english/database/db-hw/vs01.html (Accessed 14 Feb 2026).

16. Japanese Esophageal Society. Japanese Guidelines for the Diagnosis and Treatment of Esophageal Cancer 2022. Kanehara & Co. (2022).

17. Ishihara, R., Goda, K. & Oyama, T. Endoscopic diagnosis and treatment of esophageal adenocarcinoma: Introduction of Japan Esophageal Society classification of Barrett’s esophagus. J. Gastroenterol. 54, 1–9 (2019). 10.1007/s00535-018-1491-x

18. Guidozzi, N. et al. The role of artificial intelligence in the endoscopic diagnosis of esophageal cancer: A systematic review and meta-analysis. Dis. Esophagus 36, doad048 (2023). 10.1093/dote/doad048

19. Sijben, J. et al. The public’s intended uptake of hypothetical esophageal adenocarcinoma screening scenarios: A nationwide survey. Am. J. Gastroenterol. 119, 1802–1812 (2024). 10.14309/ajg.0000000000002812

20. Igakutsushin-sya. National Fee Schedule and Medical Insurance Reimbursement Table in Japan. Igakutsushin-sya (2025).

21. Bulamu, N. B. et al. Health-related quality of life associated with Barrett’s esophagus and cancer. World J. Surg. 43, 1554–1562 (2019). 10.1007/s00268-019-04936-w

22. Hur, C., Wittenberg, E., Nishioka, N. S. & Gazelle, G. S. Quality of life in patients with various Barrett’s esophagus associated health states. Health Qual. Life Outcomes 4, 45 (2006). 10.1186/1477-7525-4-45

23. Baltussen, R. M. P. M., et al. Making Choices in Health: WHO Guide to Cost-Effectiveness Analysis. World Health Organization (2003). https://apps.who.int/iris/handle/10665/42699 (Accessed 14 Feb 2026).

24. Organisation for Economic Co-operation and Development (OECD). Purchasing power parities and exchange rates. https://data.oecd.org/conversion/purchasing-power-parities-ppp.htm (Accessed 14 Feb 2026).

25. Ministry of Health, Labour and Welfare Japan. Establishment of reference values for cost-effectiveness evaluation 2018. https://www.mhlw.go.jp/file/05-Shingikai-12404000-Hokenkyoku-Iryouka/0000211609.pdf (Accessed 14 Feb 2026).

26. Husereau, D. et al. Consolidated Health Economic Evaluation Reporting Standards 2022 (CHEERS 2022) statement: Updated reporting guidance for health economic evaluations. Value Health 25, 3–9 (2022). 10.1016/j.jval.2021.11.1351

27. Kastelein, F. et al. Surveillance in patients with long-segment Barrett’s oesophagus: A cost-effectiveness analysis. Gut 64, 864–871 (2015). 10.1136/gutjnl-2014-307197

28. Vissapragada, R. et al. Evaluating cost-effectiveness of 85 endoscopic surveillance strategies of nondysplastic Barrett’s esophagus. J. Gastroenterol. Hepatol. (2026). 10.1111/jgh.70238

29. Antonios, K. et al. Risk factors for the development of Barrett’s esophagus and esophageal adenocarcinoma: A systematic review and meta-analysis. Cancer Rep. 8, e70168 (2025). 10.1002/cnr2.70168

30. Thrift, A. P. & Kendall, B. J. Epidemiology of Barrett’s esophagus and esophageal adenocarcinoma. Gastrointest. Endosc. Clin. N. Am. 36, 49–68 (2026). 10.1016/j.giec.2025.05.006

