## Supplementary Material for "Optimal Surveillance Strategies for Barrett’s Esophagus Based on Segment Length and Dysplasia Grade: A Cost-Effectiveness Study"

**Supplementary Table of Contents**

### Tables

Table S1. Base-case results and efficiency frontier (USSBE cohort)

Table S2. Base-case results and efficiency frontier (SSBE cohort)

Table S3. Base-case results and efficiency frontier (LSBE-NDBE cohort)

Table S4. Base-case results and efficiency frontier (LSBE-LGD cohort)

Table S5. Base-case results and efficiency frontier (LSBE-HGD cohort)

Table S6. Scenario results and efficiency frontier (USSBE cohort)

Table S7. Scenario results and efficiency frontier (SSBE cohort)

Table S8. Scenario results and efficiency frontier (LSBE-NDBE cohort)

Table S9. Scenario results and efficiency frontier (LSBE-LGD cohort)

Table S10. Scenario results and efficiency frontier (LSBE-HGD cohort)

### Figures

Figure S1. Tornado diagram of incremental NMB for LSBE-LGD: No surveillance vs annual endoscopy

Figure S2. Tornado diagram of incremental NMB for LSBE-HGD: Annual endoscopy vs annual breath test

Figure S3. Tornado diagram of incremental NMB for LSBE-HGD: Annual breath test vs annual miRNA test

Figure S4. Cost-effectiveness acceptability curve for LSBE-LGD including no surveillance

Figure S5. Cost-effectiveness acceptability curve for LSBE-HGD including no surveillance

Figure S6. EAC deaths averted per 10,000 LSBE-LGD patients over 10 years and over a lifetime

Figure S7. EAC deaths averted per 10,000 LSBE-HGD patients over 10 years and over a lifetime

Figure S8. Additional costs per EAC death averted among 10,000 LSBE-LGD patients over 10 years and over a lifetime

Figure S9. Additional costs per EAC death averted among 10,000 LSBE-HGD patients over 10 years and over a lifetime

**General caption for Tables S1-S10**

All tables report total costs, total QALYs, ICERs, and NMBs for the evaluated screening strategies. All monetary values are presented in US$.

**General caption for Figures S1-S3**

Each tornado diagram shows how variation in individual parameters across their plausible ranges influences the incremental NMB between the compared screening strategies. Wider bars indicate greater impact on cost-effectiveness results.

**General caption for Figures S6-S9**

All values represent differences compared with no surveillance. Each figure presents results per 10,000 patients for 10-year and lifetime horizons.

**List of Abbreviations**

AI: Artificial intelligence

BE: Barrett's esophagus

EAC: Esophageal adenocarcinoma

HGD: High-grade dysplasia

ICER: Incremental cost-effectiveness ratio

LGD: Low-grade dysplasia

LSBE: Long-segment Barrett's esophagus

miRNA: MicroRNA

NDBE: Nondysplastic Barrett's esophagus

NMB: Net monetary benefit

QALY: Quality-adjusted life-year

SSBE: Short-segment Barrett's esophagus

USSBE: Ultrashort-segment Barrett's esophagus

**Table S1.** Base-case results and efficiency frontier (USSBE cohort)

| Strategy | Cost (US$) | Incremental Cost (US$) | Effectiveness (QALY) | Incremental Effectiveness (QALY) | ICER (US$/QALY) | NMB (US$) |
|---|---|---|---|---|---|---|
| No surveillance | 28 | Ref | 17.39255 | Ref | Ref | 869,600 |
| Endoscopy every 10 years | 588 | 561 | 17.39258 | 0.00003 | Extendedly dominated | 869,041 |
| Endoscopy every 5 years | 959 | 371 | 17.39261 | 0.00002 | Extendedly dominated | 868,671 |
| Breath test every 10 years | 1,002 | 43 | 17.39259 | − 0.00002 | Absolutely dominated | 868,628 |
| Endoscopy every 4 years | 1,145 | 186 | 17.39262 | 0.00001 | Extendedly dominated | 868,486 |
| Endoscopy every 3 years | 1,456 | 310 | 17.39264 | 0.00002 | Extendedly dominated | 868,176 |
| Sponge test every 10 years | 1,491 | 36 | 17.39258 | − 0.00006 | Absolutely dominated | 868,138 |
| Breath test every 5 years | 1,646 | 190 | 17.39261 | − 0.00003 | Absolutely dominated | 867,985 |
| Breath test every 4 years | 1,969 | 514 | 17.39263 | − 0.00001 | Absolutely dominated | 867,662 |
| Endoscopy every 2 years | 2,077 | 621 | 17.39268 | 0.00004 | Extendedly dominated | 867,557 |
| Sponge test every 5 years | 2,459 | 382 | 17.39259 | − 0.00008 | Absolutely dominated | 867,170 |
| Breath test every 3 years | 2,509 | 432 | 17.39265 | − 0.00003 | Absolutely dominated | 867,124 |
| miRNA test every 10 years | 2,757 | 680 | 17.39259 | − 0.00009 | Absolutely dominated | 866,872 |
| Sponge test every 4 years | 2,945 | 868 | 17.39260 | − 0.00007 | Absolutely dominated | 866,685 |
| Breath test every 2 years | 3,588 | 1,511 | 17.39269 | 0.00001 | Extendedly dominated | 866,046 |
| Sponge test every 3 years | 3,756 | 168 | 17.39262 | − 0.00007 | Absolutely dominated | 865,875 |
| Annual endoscopy | 3,942 | 3,915 | 17.39280 | 0.00024 | 16,021,510 | 865,697 |
| miRNA test every 5 years | 4,563 | 620 | 17.39262 | − 0.00018 | Absolutely dominated | 865,068 |
| Sponge test every 2 years | 5,379 | 1,436 | 17.39265 | − 0.00015 | Absolutely dominated | 864,254 |
| miRNA test every 4 years | 5,469 | 1,527 | 17.39263 | − 0.00017 | Absolutely dominated | 864,162 |
| Annual breath test | 6,830 | 2,887 | 17.39282 | 0.00003 | 109,073,360 | 862,812 |
| miRNA test every 3 years | 6,981 | 152 | 17.39265 | − 0.00017 | Absolutely dominated | 862,651 |
| miRNA test every 2 years | 10,008 | 3,178 | 17.39270 | − 0.00012 | Absolutely dominated | 859,627 |
| Annual sponge test | 10,249 | 3,420 | 17.39274 | − 0.00008 | Absolutely dominated | 859,388 |
| Annual miRNA test | 19,092 | 12,263 | 17.39284 | 0.00001 | 858,476,343 | 850,549 |

| Strategy | Cost (US$) | Incremental Cost (US$) | Effectiveness (QALY) | Incremental Effectiveness (QALY) | ICER (US$/QALY) | NMB (US$) |
|---|---|---|---|---|---|---|
| No surveillance | 28 | Ref | 17.39255 | Ref | Ref | 869,600 |
| Annual endoscopy | 3,942 | 3,915 | 17.39280 | 0.00024 | 16,021,510 | 865,697 |
| Annual breath test | 6,830 | 2,887 | 17.39282 | 0.00003 | 109,073,360 | 862,812 |
| Annual miRNA test | 19,092 | 12,263 | 17.39284 | 0.00001 | 858,476,343 | 850,549 |

Dominance: “Absolutely dominated” indicates higher cost and lower effectiveness. “Extendedly dominated” indicates a higher ICER than the next more effective strategy. All costs and benefits are discounted at 3% annually. Willingness-to-pay threshold was set at US$50,000 per QALY. Ref indicates the reference (baseline) strategy.

**Table S2.** Base-case results and efficiency frontier (SSBE cohort)

| Strategy | Cost (US$) | Incremental Cost (US$) | Effectiveness (QALY) | Incremental Effectiveness (QALY) | ICER (US$/QALY) | NMB (US$) |
|---|---|---|---|---|---|---|
| No surveillance | 225 | Ref | 19.00488 | Ref | Ref | 950,019 |
| Endoscopy every 10 years | 783 | 558 | 19.00516 | 0.00028 | 2,004,493 | 949,475 |
| Endoscopy every 5 years | 1,151 | 368 | 19.00536 | 0.00020 | 1,838,954 | 949,117 |
| Breath test every 10 years | 1,195 | 44 | 19.00519 | −0.00017 | Absolutely dominated | 949,064 |
| Endoscopy every 4 years | 1,336 | 185 | 19.00546 | 0.00010 | Extendedly dominated | 948,937 |
| Endoscopy every 3 years | 1,645 | 309 | 19.00563 | 0.00017 | Extendedly dominated | 948,637 |
| Sponge test every 10 years | 1,684 | 39 | 19.00510 | −0.00053 | Absolutely dominated | 948,571 |
| Breath test every 5 years | 1,836 | 191 | 19.00541 | −0.00022 | Absolutely dominated | 948,435 |
| Breath test every 4 years | 2,158 | 513 | 19.00552 | −0.00011 | Absolutely dominated | 948,118 |
| Endoscopy every 2 years | 2,262 | 618 | 19.00597 | 0.00034 | Extendedly dominated | 948,036 |
| Sponge test every 5 years | 2,648 | 386 | 19.00525 | −0.00071 | Absolutely dominated | 947,615 |
| Breath test every 3 years | 2,694 | 432 | 19.00571 | −0.00026 | Absolutely dominated | 947,591 |
| miRNA test every 10 years | 2,947 | 684 | 19.00520 | −0.00076 | Absolutely dominated | 947,313 |
| Sponge test every 4 years | 3,132 | 869 | 19.00533 | −0.00064 | Absolutely dominated | 947,135 |
| Breath test every 2 years | 3,769 | 1,506 | 19.00608 | 0.00012 | Extendedly dominated | 946,535 |
| Sponge test every 3 years | 3,939 | 171 | 19.00546 | −0.00062 | Absolutely dominated | 946,334 |
| Annual endoscopy | 4,117 | 3,892 | 19.00698 | 0.00210 | 1,851,435 | 946,232 |
| miRNA test every 5 years | 4,745 | 628 | 19.00544 | −0.00154 | Absolutely dominated | 945,527 |
| Sponge test every 2 years | 5,555 | 1,439 | 19.00572 | −0.00126 | Absolutely dominated | 944,731 |
| miRNA test every 4 years | 5,648 | 1,531 | 19.00556 | −0.00143 | Absolutely dominated | 944,630 |
| Annual breath test | 6,994 | 2,877 | 19.00721 | 0.00023 | 12,632,720 | 943,367 |
| miRNA test every 3 years | 7,154 | 160 | 19.00575 | −0.00146 | Absolutely dominated | 943,134 |
| miRNA test every 2 years | 10,168 | 3,175 | 19.00615 | −0.00106 | Absolutely dominated | 940,139 |
| Annual sponge test | 10,406 | 3,412 | 19.00652 | −0.00069 | Absolutely dominated | 939,920 |
| Annual miRNA test | 19,217 | 12,223 | 19.00733 | 0.00012 | 99,460,943 | 931,150 |

| Strategy | Cost (US$) | Incremental Cost (US$) | Effectiveness (QALY) | Incremental Effectiveness (QALY) | ICER (US$/QALY) | NMB (US$) |
|---|---|---|---|---|---|---|
| No surveillance | 225 | Ref | 19.00488 | Ref | Ref | 950,019 |
| Annual endoscopy | 4,117 | 3,892 | 19.00698 | 0.00210 | 1,851,435 | 946,232 |
| Annual breath test | 6,994 | 2,877 | 19.00721 | 0.00023 | 12,632,720 | 943,367 |
| Annual miRNA test | 19,217 | 12,223 | 19.00733 | 0.00012 | 99,460,943 | 931,150 |

Same definitions as Table S1.

**Table S3.** Base-case results and efficiency frontier (LSBE-NDBE cohort)

| Strategy | Cost (US$) | Incremental Cost (US$) | Effectiveness (QALY) | Incremental Effectiveness (QALY) | ICER (US$/QALY) | NMB (US$) |
|---|---|---|---|---|---|---|
| No surveillance | 1,770 | Ref | 18.81182 | Ref | Ref | 938,821 |
| Endoscopy every 10 years | 2,309 | 539 | 18.81403 | 0.00222 | Extendedly dominated | 938,393 |
| Endoscopy every 5 years | 2,659 | 351 | 18.81562 | 0.00158 | Extendedly dominated | 938,121 |
| Breath test every 10 years | 2,713 | 53 | 18.81427 | −0.00134 | Absolutely dominated | 938,001 |
| Endoscopy every 4 years | 2,836 | 176 | 18.81642 | 0.00080 | Extendedly dominated | 937,985 |
| Endoscopy every 3 years | 3,130 | 294 | 18.81775 | 0.00134 | Extendedly dominated | 937,758 |
| Sponge test every 10 years | 3,197 | 67 | 18.81354 | −0.00421 | Absolutely dominated | 937,480 |
| Breath test every 5 years | 3,328 | 198 | 18.81603 | −0.00172 | Absolutely dominated | 937,473 |
| Breath test every 4 years | 3,637 | 507 | 18.81691 | −0.00084 | Absolutely dominated | 937,209 |
| Endoscopy every 2 years | 3,718 | 588 | 18.82043 | 0.00267 | Extendedly dominated | 937,303 |
| Sponge test every 5 years | 4,129 | 411 | 18.81478 | −0.00565 | Absolutely dominated | 936,610 |
| Breath test every 3 years | 4,152 | 434 | 18.81839 | −0.00203 | Absolutely dominated | 936,768 |
| miRNA test every 10 years | 4,434 | 717 | 18.81440 | −0.00602 | Absolutely dominated | 936,286 |
| Sponge test every 4 years | 4,597 | 879 | 18.81540 | −0.00503 | Absolutely dominated | 936,173 |
| Breath test every 2 years | 5,184 | 1,466 | 18.82136 | 0.00093 | Extendedly dominated | 935,884 |
| Sponge test every 3 years | 5,378 | 194 | 18.81644 | −0.00492 | Absolutely dominated | 935,444 |
| Annual endoscopy | 5,484 | 3,714 | 18.82846 | 0.01664 | 223,133 | 935,939 |
| miRNA test every 5 years | 6,175 | 691 | 18.81625 | −0.01221 | Absolutely dominated | 934,637 |
| Sponge test every 2 years | 6,941 | 1,457 | 18.81852 | −0.00994 | Absolutely dominated | 933,985 |
| miRNA test every 4 years | 7,049 | 1,565 | 18.81718 | −0.01128 | Absolutely dominated | 933,810 |
| Annual breath test | 8,280 | 2,796 | 18.83026 | 0.00180 | 1,550,676 | 933,233 |
| miRNA test every 3 years | 8,508 | 228 | 18.81874 | −0.01152 | Absolutely dominated | 932,429 |
| miRNA test every 2 years | 11,427 | 3,148 | 18.82186 | −0.00840 | Absolutely dominated | 929,666 |
| Annual sponge test | 11,633 | 3,353 | 18.82478 | −0.00548 | Absolutely dominated | 929,606 |
| Annual miRNA test | 20,191 | 11,912 | 18.83123 | 0.00097 | 12,242,182 | 921,370 |

| Strategy | Cost (US$) | Incremental Cost (US$) | Effectiveness (QALY) | Incremental Effectiveness (QALY) | ICER (US$/QALY) | NMB (US$) |
|---|---|---|---|---|---|---|
| No surveillance | 1,770 | Ref | 18.81182 | Ref | Ref | 938,821 |
| Annual endoscopy | 5,484 | 3,714 | 18.82846 | 0.01664 | 223,133 | 935,939 |
| Annual breath test | 8,280 | 2,796 | 18.83026 | 0.00180 | 1,550,676 | 933,233 |
| Annual miRNA test | 20,191 | 11,912 | 18.83123 | 0.00097 | 12,242,182 | 921,370 |

Same definitions as Table S1.

**Table S4.** Base-case results and efficiency frontier (LSBE-LGD cohort)

| Strategy | Cost (US$) | Incremental Cost (US$) | Effectiveness (QALY) | Incremental Effectiveness (QALY) | ICER (US$/QALY) | NMB (US$) |
|---|---|---|---|---|---|---|
| No surveillance | 8,655 | Ref | 16.99090 | Ref | Ref | 840,890 |
| Endoscopy every 10 years | 9,104 | 449 | 17.00190 | 0.01099 | Extendedly dominated | 840,991 |
| Endoscopy every 5 years | 9,374 | 271 | 17.00946 | 0.00757 | Extendedly dominated | 841,099 |
| Breath test every 10 years | 9,471 | 96 | 17.00309 | −0.00638 | Absolutely dominated | 840,684 |
| Endoscopy every 4 years | 9,511 | 136 | 17.01329 | 0.00383 | Extendedly dominated | 841,154 |
| Endoscopy every 3 years | 9,738 | 227 | 17.01970 | 0.00640 | Extendedly dominated | 841,247 |
| Sponge test every 10 years | 9,932 | 194 | 16.99946 | −0.02023 | Absolutely dominated | 840,041 |
| Breath test every 5 years | 9,968 | 230 | 17.01148 | −0.00822 | Absolutely dominated | 840,605 |
| Endoscopy every 2 years | 10,194 | 456 | 17.03253 | 0.01283 | Extendedly dominated | 841,433 |
| Breath test every 4 years | 10,219 | 25 | 17.01572 | −0.01681 | Absolutely dominated | 840,567 |
| Breath test every 3 years | 10,637 | 443 | 17.02281 | −0.00971 | Absolutely dominated | 840,504 |
| Sponge test every 5 years | 10,718 | 524 | 17.00536 | −0.02717 | Absolutely dominated | 839,550 |
| miRNA test every 10 years | 11,053 | 860 | 17.00373 | −0.02880 | Absolutely dominated | 839,133 |
| Sponge test every 4 years | 11,113 | 920 | 17.00834 | −0.02419 | Absolutely dominated | 839,304 |
| Breath test every 2 years | 11,474 | 1,280 | 17.03704 | 0.00451 | Extendedly dominated | 840,378 |
| Annual endoscopy | 11,561 | 2,907 | 17.07109 | 0.08018 | 36,250 | 841,993 |
| Sponge test every 3 years | 11,774 | 212 | 17.01333 | −0.05776 | Absolutely dominated | 838,893 |
| miRNA test every 5 years | 12,531 | 970 | 17.01256 | −0.05853 | Absolutely dominated | 838,097 |
| Sponge test every 2 years | 13,096 | 1,534 | 17.02332 | −0.04776 | Absolutely dominated | 838,070 |
| miRNA test every 4 years | 13,275 | 1,714 | 17.01703 | −0.05406 | Absolutely dominated | 837,576 |
| Annual breath test | 13,987 | 2,426 | 17.07977 | 0.00869 | 279,252 | 840,002 |
| miRNA test every 3 years | 14,517 | 530 | 17.02450 | −0.05528 | Absolutely dominated | 836,708 |
| miRNA test every 2 years | 17,004 | 3,017 | 17.03947 | −0.04030 | Absolutely dominated | 834,970 |
| Annual sponge test | 17,066 | 3,078 | 17.05335 | −0.02642 | Absolutely dominated | 835,602 |
| Annual miRNA test | 24,470 | 10,483 | 17.08446 | 0.00469 | 2,236,293 | 829,753 |

| Strategy | Cost (US$) | Incremental Cost (US$) | Effectiveness (QALY) | Incremental Effectiveness (QALY) | ICER (US$/QALY) | NMB (US$) |
|---|---|---|---|---|---|---|
| No surveillance | 8,655 | Ref | 16.99090 | Ref | Ref | 840,890 |
| Annual endoscopy | 11,561 | 2,907 | 17.07109 | 0.08018 | 36,250 | 841,993 |
| Annual breath test | 13,987 | 2,426 | 17.07977 | 0.00869 | 279,252 | 840,002 |
| Annual miRNA test | 24,470 | 10,483 | 17.08446 | 0.00469 | 2,236,293 | 829,753 |

Same definitions as Table S1.

**Table S5.** Base-case results and efficiency frontier (LSBE-HGD cohort)

| Strategy | Cost (US$) | Incremental Cost (US$) | Effectiveness (QALY) | Incremental Effectiveness (QALY) | ICER (US$/QALY) | NMB (US$) |
|---|---|---|---|---|---|---|
| Annual endoscopy | 35,165 | Ref | 12.63758 | Ref | Ref | 596,713 |
| Endoscopy every 2 years | 35,522 | 357 | 12.47124 | −0.16634 | Absolutely dominated | 588,040 |
| Endoscopy every 3 years | 35,640 | 474 | 12.41655 | −0.22103 | Absolutely dominated | 585,188 |
| Endoscopy every 4 years | 35,697 | 532 | 12.38976 | −0.24782 | Absolutely dominated | 583,791 |
| Endoscopy every 5 years | 35,731 | 566 | 12.37412 | −0.26346 | Absolutely dominated | 582,975 |
| Endoscopy every 10 years | 35,793 | 628 | 12.34581 | −0.29177 | Absolutely dominated | 581,497 |
| Annual breath test | 35,821 | 655 | 12.67773 | 0.04015 | 16,318 | 598,066 |
| No surveillance | 35,862 | 41 | 12.26694 | −0.41079 | Absolutely dominated | 577,485 |
| Breath test every 2 years | 35,911 | 90 | 12.49337 | −0.18436 | Absolutely dominated | 588,758 |
| Breath test every 3 years | 35,940 | 120 | 12.43275 | −0.24498 | Absolutely dominated | 585,697 |
| Breath test every 4 years | 35,955 | 134 | 12.40306 | −0.27467 | Absolutely dominated | 584,198 |
| Breath test every 5 years | 35,963 | 143 | 12.38573 | −0.29200 | Absolutely dominated | 583,323 |
| Breath test every 10 years | 35,979 | 158 | 12.35436 | −0.32338 | Absolutely dominated | 581,739 |
| Sponge test every 10 years | 36,370 | 550 | 12.32837 | −0.34936 | Absolutely dominated | 580,048 |
| Sponge test every 5 years | 36,477 | 657 | 12.35042 | −0.32731 | Absolutely dominated | 581,044 |
| Sponge test every 4 years | 36,535 | 715 | 12.36260 | −0.31513 | Absolutely dominated | 581,594 |
| Sponge test every 3 years | 36,634 | 814 | 12.38346 | −0.29427 | Absolutely dominated | 582,539 |
| Sponge test every 2 years | 36,836 | 1,015 | 12.42606 | −0.25167 | Absolutely dominated | 584,467 |
| miRNA test every 10 years | 36,917 | 1,097 | 12.35897 | −0.31876 | Absolutely dominated | 581,031 |
| miRNA test every 5 years | 37,159 | 1,338 | 12.39200 | −0.28573 | Absolutely dominated | 582,441 |
| miRNA test every 4 years | 37,290 | 1,470 | 12.41024 | −0.26749 | Absolutely dominated | 583,222 |
| Annual sponge test | 37,447 | 1,627 | 12.55561 | −0.12212 | Absolutely dominated | 590,333 |
| miRNA test every 3 years | 37,514 | 1,693 | 12.44150 | −0.23623 | Absolutely dominated | 584,561 |
| miRNA test every 2 years | 37,969 | 2,148 | 12.50531 | −0.17242 | Absolutely dominated | 587,297 |
| Annual miRNA test | 39,350 | 3,529 | 12.69940 | 0.02167 | 162,872 | 595,620 |

| Strategy | Cost (US$) | Incremental Cost (US$) | Effectiveness (QALY) | Incremental Effectiveness (QALY) | ICER (US$/QALY) | NMB (US$) |
|---|---|---|---|---|---|---|
| Annual endoscopy | 35,165 | Ref | 12.63758 | Ref | Ref | 596,713 |
| Annual breath test | 35,821 | 655 | 12.67773 | 0.04015 | 16,318 | 598,066 |
| Annual miRNA test | 39,350 | 3,529 | 12.69940 | 0.02167 | 162,872 | 595,620 |

Same definitions as Table S1.

**Table S6.** Scenario results and efficiency frontier (USSBE cohort)

| Strategy | Cost (US$) | Incremental Cost (US$) | Effectiveness (QALY) | Incremental Effectiveness (QALY) | ICER (US$/QALY) | NMB (US$) |
|---|---|---|---|---|---|---|
| No surveillance | 28 | Ref | 19.02934 | Ref | Ref | 951,439 |
| AI-assisted endoscopy every 10 years | 641 | 613 | 19.02939 | 0.00004 | Extendedly dominated | 950,828 |
| AI-assisted endoscopy every 5 years | 1,046 | 406 | 19.02942 | 0.00003 | Extendedly dominated | 950,424 |
| Breath test every 10 years | 1,077 | 30 | 19.02939 | −0.00003 | Absolutely dominated | 950,393 |
| AI-assisted endoscopy every 4 years | 1,250 | 204 | 19.02943 | 0.00002 | Extendedly dominated | 950,221 |
| Sponge test every 10 years | 1,538 | 288 | 19.02938 | −0.00006 | Absolutely dominated | 949,931 |
| AI-assisted endoscopy every 3 years | 1,590 | 340 | 19.02946 | 0.00003 | Extendedly dominated | 949,883 |
| Breath test every 5 years | 1,771 | 181 | 19.02942 | −0.00003 | Absolutely dominated | 949,701 |
| Breath test every 4 years | 2,119 | 529 | 19.02944 | −0.00002 | Absolutely dominated | 949,353 |
| AI-assisted endoscopy every 2 years | 2,270 | 680 | 19.02951 | 0.00005 | Extendedly dominated | 949,206 |
| Sponge test every 5 years | 2,537 | 267 | 19.02940 | −0.00011 | Absolutely dominated | 948,933 |
| Breath test every 3 years | 2,700 | 430 | 19.02947 | −0.00004 | Absolutely dominated | 948,774 |
| miRNA test every 10 years | 2,822 | 552 | 19.02939 | −0.00012 | Absolutely dominated | 948,648 |
| Sponge test every 4 years | 3,038 | 769 | 19.02941 | −0.00010 | Absolutely dominated | 948,432 |
| Breath test every 2 years | 3,863 | 1,593 | 19.02953 | 0.00002 | Extendedly dominated | 947,613 |
| Sponge test every 3 years | 3,875 | 12 | 19.02943 | −0.00010 | Absolutely dominated | 947,597 |
| Annual AI-assisted endoscopy | 4,310 | 4,283 | 19.02966 | 0.00032 | 13,315,317 | 947,173 |
| miRNA test every 5 years | 4,670 | 360 | 19.02943 | −0.00024 | Absolutely dominated | 946,801 |
| Sponge test every 2 years | 5,549 | 1,239 | 19.02947 | −0.00019 | Absolutely dominated | 945,925 |
| miRNA test every 4 years | 5,598 | 1,288 | 19.02945 | −0.00022 | Absolutely dominated | 945,874 |
| miRNA test every 3 years | 7,146 | 2,836 | 19.02948 | −0.00019 | Absolutely dominated | 944,328 |
| Annual breath test | 7,354 | 3,044 | 19.02970 | 0.00003 | 87,353,552 | 944,131 |
| miRNA test every 2 years | 10,244 | 2,890 | 19.02954 | −0.00016 | Absolutely dominated | 941,233 |
| Annual sponge test | 10,575 | 3,221 | 19.02959 | −0.00011 | Absolutely dominated | 940,905 |
| Annual miRNA test | 19,543 | 12,189 | 19.02972 | 0.00002 | 648,276,885 | 931,943 |

| Strategy | Cost (US$) | Incremental Cost (US$) | Effectiveness (QALY) | Incremental Effectiveness (QALY) | ICER (US$/QALY) | NMB (US$) |
|---|---|---|---|---|---|---|
| No surveillance | 28 | Ref | 19.02934 | Ref | Ref | 951,439 |
| Annual AI-assisted endoscopy | 4,310 | 4,283 | 19.02966 | 0.00032 | 13,315,317 | 947,173 |
| Annual breath test | 7,354 | 3,044 | 19.02970 | 0.00003 | 87,353,552 | 944,131 |
| Annual miRNA test | 19,543 | 12,189 | 19.02972 | 0.00002 | 648,276,885 | 931,943 |

Same definitions as Table S1.

**Table S7.** Scenario results and efficiency frontier (SSBE cohort)

| Strategy | Cost (US$) | Incremental Cost (US$) | Effectiveness (QALY) | Incremental Effectiveness (QALY) | ICER (US$/QALY) | NMB (US$) |
|---|---|---|---|---|---|---|
| No surveillance | 225 | Ref | 19.00488 | Ref | Ref | 950,019 |
| AI-assisted endoscopy every 10 years | 835 | 610 | 19.00522 | 0.00035 | Extendedly dominated | 949,426 |
| AI-assisted endoscopy every 5 years | 1,238 | 403 | 19.00547 | 0.00025 | Extendedly dominated | 949,036 |
| Breath test every 10 years | 1,270 | 32 | 19.00526 | −0.00021 | Absolutely dominated | 948,993 |
| AI-assisted endoscopy every 4 years | 1,440 | 202 | 19.00560 | 0.00013 | Extendedly dominated | 948,840 |
| Sponge test every 10 years | 1,730 | 290 | 19.00515 | −0.00045 | Absolutely dominated | 948,527 |
| AI-assisted endoscopy every 3 years | 1,778 | 337 | 19.00581 | 0.00021 | Extendedly dominated | 948,513 |
| Breath test every 5 years | 1,960 | 182 | 19.00554 | −0.00027 | Absolutely dominated | 948,317 |
| Breath test every 4 years | 2,306 | 528 | 19.00568 | −0.00013 | Absolutely dominated | 947,978 |
| AI-assisted endoscopy every 2 years | 2,453 | 676 | 19.00623 | 0.00042 | Extendedly dominated | 947,858 |
| Sponge test every 5 years | 2,725 | 272 | 19.00534 | −0.00089 | Absolutely dominated | 947,542 |
| Breath test every 3 years | 2,884 | 431 | 19.00591 | −0.00032 | Absolutely dominated | 947,411 |
| miRNA test every 10 years | 3,011 | 558 | 19.00528 | −0.00094 | Absolutely dominated | 947,253 |
| Sponge test every 4 years | 3,224 | 771 | 19.00544 | −0.00079 | Absolutely dominated | 947,048 |
| Breath test every 2 years | 4,041 | 1,588 | 19.00637 | 0.00015 | Extendedly dominated | 946,278 |
| Sponge test every 3 years | 4,057 | 16 | 19.00560 | −0.00077 | Absolutely dominated | 946,223 |
| Annual AI-assisted endoscopy | 4,481 | 4,256 | 19.00749 | 0.00261 | 1,632,816 | 945,893 |
| miRNA test every 5 years | 4,851 | 371 | 19.00557 | −0.00191 | Absolutely dominated | 945,427 |
| Sponge test every 2 years | 5,724 | 1,243 | 19.00593 | −0.00156 | Absolutely dominated | 944,572 |
| miRNA test every 4 years | 5,775 | 1,295 | 19.00572 | −0.00177 | Absolutely dominated | 944,510 |
| miRNA test every 3 years | 7,317 | 2,836 | 19.00596 | −0.00152 | Absolutely dominated | 942,981 |
| Annual breath test | 7,513 | 3,033 | 19.00777 | 0.00028 | 10,738,961 | 942,875 |
| miRNA test every 2 years | 10,402 | 2,889 | 19.00645 | −0.00132 | Absolutely dominated | 939,920 |
| Annual sponge test | 10,728 | 3,215 | 19.00691 | −0.00086 | Absolutely dominated | 939,617 |
| Annual miRNA test | 19,663 | 12,150 | 19.00792 | 0.00015 | 79,728,307 | 930,733 |

| Strategy | Cost (US$) | Incremental Cost (US$) | Effectiveness (QALY) | Incremental Effectiveness (QALY) | ICER (US$/QALY) | NMB (US$) |
|---|---|---|---|---|---|---|
| No surveillance | 225 | Ref | 19.00488 | Ref | Ref | 950,019 |
| Annual AI-assisted endoscopy | 4,481 | 4,256 | 19.00749 | 0.00261 | 1,632,816 | 945,893 |
| Annual breath test | 7,513 | 3,033 | 19.00777 | 0.00028 | 10,738,961 | 942,875 |
| Annual miRNA test | 19,663 | 12,150 | 19.00792 | 0.00015 | 79,728,307 | 930,733 |

Same definitions as Table S1.

**Table S8.** Scenario results and efficiency frontier (LSBE-NDBE cohort)

| Strategy | Cost (US$) | Incremental Cost (US$) | Effectiveness (QALY) | Incremental Effectiveness (QALY) | ICER (US$/QALY) | NMB (US$) |
|---|---|---|---|---|---|---|
| No surveillance | 1,770 | Ref | 18.81182 | Ref | Ref | 938,821 |
| AI-assisted endoscopy every 10 years | 2,358 | 587 | 18.81456 | 0.00275 | Extendedly dominated | 938,371 |
| AI-assisted endoscopy every 5 years | 2,740 | 382 | 18.81653 | 0.00196 | Extendedly dominated | 938,086 |
| Breath test every 10 years | 2,784 | 43 | 18.81486 | −0.00167 | Absolutely dominated | 937,960 |
| AI-assisted endoscopy every 4 years | 2,932 | 192 | 18.81752 | 0.00099 | Extendedly dominated | 937,944 |
| Sponge test every 10 years | 3,240 | 308 | 18.81396 | −0.00356 | Absolutely dominated | 937,457 |
| AI-assisted endoscopy every 3 years | 3,253 | 320 | 18.81917 | 0.00166 | Extendedly dominated | 937,706 |
| Breath test every 5 years | 3,444 | 192 | 18.81704 | −0.00214 | Absolutely dominated | 937,408 |
| Breath test every 4 years | 3,776 | 524 | 18.81814 | −0.00104 | Absolutely dominated | 937,131 |
| AI-assisted endoscopy every 2 years | 3,894 | 641 | 18.82249 | 0.00332 | Extendedly dominated | 937,230 |
| Sponge test every 5 years | 4,201 | 307 | 18.81549 | −0.00701 | Absolutely dominated | 936,574 |
| Breath test every 3 years | 4,330 | 436 | 18.81997 | −0.00252 | Absolutely dominated | 936,669 |
| miRNA test every 10 years | 4,495 | 600 | 18.81502 | −0.00747 | Absolutely dominated | 936,257 |
| Sponge test every 4 years | 4,683 | 789 | 18.81626 | −0.00623 | Absolutely dominated | 936,130 |
| Breath test every 2 years | 5,438 | 1,544 | 18.82365 | 0.00116 | Extendedly dominated | 935,744 |
| Sponge test every 3 years | 5,487 | 49 | 18.81755 | −0.00610 | Absolutely dominated | 935,390 |
| Annual AI-assisted endoscopy | 5,820 | 4,049 | 18.83245 | 0.02064 | 196,213 | 935,803 |
| miRNA test every 5 years | 6,274 | 455 | 18.81731 | −0.01514 | Absolutely dominated | 934,592 |
| Sponge test every 2 years | 7,098 | 1,278 | 18.82013 | −0.01232 | Absolutely dominated | 933,909 |
| miRNA test every 4 years | 7,168 | 1,348 | 18.81847 | −0.01398 | Absolutely dominated | 933,756 |
| miRNA test every 3 years | 8,659 | 2,840 | 18.82040 | −0.01205 | Absolutely dominated | 932,361 |
| Annual breath test | 8,765 | 2,946 | 18.83469 | 0.00224 | 1,317,489 | 932,969 |
| miRNA test every 2 years | 11,644 | 2,879 | 18.82427 | −0.01042 | Absolutely dominated | 929,570 |
| Annual sponge test | 11,932 | 3,167 | 18.82789 | −0.00680 | Absolutely dominated | 929,462 |
| Annual miRNA test | 20,604 | 11,839 | 18.83590 | 0.00121 | 9,812,423 | 921,191 |

| Strategy | Cost (US$) | Incremental Cost (US$) | Effectiveness (QALY) | Incremental Effectiveness (QALY) | ICER (US$/QALY) | NMB (US$) |
|---|---|---|---|---|---|---|
| No surveillance | 1,770 | Ref | 18.81182 | Ref | Ref | 938,821 |
| Annual AI-assisted endoscopy | 5,820 | 4,049 | 18.83245 | 0.02064 | 196,213 | 935,803 |
| Annual breath test | 8,765 | 2,946 | 18.83469 | 0.00224 | 1,317,489 | 932,969 |
| Annual miRNA test | 20,604 | 11,839 | 18.83590 | 0.00121 | 9,812,423 | 921,191 |

Same definitions as Table S1.

**Table S9.** Scenario results and efficiency frontier (LSBE-LGD cohort)

| Strategy | Cost (US$) | Incremental Cost (US$) | Effectiveness (QALY) | Incremental Effectiveness (QALY) | ICER (US$/QALY) | NMB (US$) |
|---|---|---|---|---|---|---|
| No surveillance | 8,655 | Ref | 16.99090 | Ref | Ref | 840,890 |
| AI-assisted endoscopy every 10 years | 9,137 | 483 | 17.00453 | 0.01363 | Extendedly dominated | 841,089 |
| AI-assisted endoscopy every 5 years | 9,427 | 290 | 17.01392 | 0.00939 | Extendedly dominated | 841,269 |
| Breath test every 10 years | 9,523 | 96 | 17.00601 | −0.00791 | Absolutely dominated | 840,777 |
| AI-assisted endoscopy every 4 years | 9,573 | 146 | 17.01867 | 0.00475 | Extendedly dominated | 841,360 |
| AI-assisted endoscopy every 3 years | 9,817 | 244 | 17.02661 | 0.00794 | Extendedly dominated | 841,513 |
| Sponge test every 10 years | 9,963 | 146 | 17.00152 | −0.02509 | Absolutely dominated | 840,113 |
| Breath test every 5 years | 10,051 | 235 | 17.01641 | −0.01019 | Absolutely dominated | 840,769 |
| AI-assisted endoscopy every 2 years | 10,305 | 488 | 17.04252 | 0.01591 | Extendedly dominated | 841,821 |
| Breath test every 4 years | 10,317 | 13 | 17.02167 | −0.02084 | Absolutely dominated | 840,767 |
| Breath test every 3 years | 10,761 | 456 | 17.03047 | −0.01205 | Absolutely dominated | 840,763 |
| Sponge test every 5 years | 10,767 | 463 | 17.00883 | −0.03369 | Absolutely dominated | 839,674 |
| miRNA test every 10 years | 11,095 | 791 | 17.00681 | −0.03571 | Absolutely dominated | 839,245 |
| Sponge test every 4 years | 11,172 | 867 | 17.01253 | −0.02999 | Absolutely dominated | 839,455 |
| Breath test every 2 years | 11,649 | 1,345 | 17.04811 | 0.00559 | Extendedly dominated | 840,756 |
| Annual AI-assisted endoscopy | 11,769 | 3,114 | 17.09033 | 0.09943 | 31,323 | 842,747 |
| Sponge test every 3 years | 11,847 | 78 | 17.01871 | −0.07162 | Absolutely dominated | 839,088 |
| miRNA test every 5 years | 12,598 | 829 | 17.01776 | −0.07257 | Absolutely dominated | 838,290 |
| Sponge test every 2 years | 13,200 | 1,431 | 17.03110 | −0.05923 | Absolutely dominated | 838,355 |
| miRNA test every 4 years | 13,354 | 1,585 | 17.02330 | −0.06703 | Absolutely dominated | 837,811 |
| Annual breath test | 14,317 | 2,548 | 17.10110 | 0.01077 | 236,568 | 840,738 |
| miRNA test every 3 years | 14,616 | 299 | 17.03256 | −0.06854 | Absolutely dominated | 837,012 |
| miRNA test every 2 years | 17,143 | 2,826 | 17.05113 | −0.04997 | Absolutely dominated | 835,413 |
| Annual sponge test | 17,261 | 2,943 | 17.06834 | −0.03276 | Absolutely dominated | 836,156 |
| Annual miRNA test | 24,731 | 10,414 | 17.10691 | 0.00581 | 1,791,535 | 830,615 |

| Strategy | Cost (US$) | Incremental Cost (US$) | Effectiveness (QALY) | Incremental Effectiveness (QALY) | ICER (US$/QALY) | NMB (US$) |
|---|---|---|---|---|---|---|
| No surveillance | 8,655 | Ref | 16.99090 | Ref | Ref | 840,890 |
| Annual AI-assisted endoscopy | 11,769 | 3,114 | 17.09033 | 0.09943 | 31,323 | 842,747 |
| Annual breath test | 14,317 | 2,548 | 17.10110 | 0.01077 | 236,568 | 840,738 |
| Annual miRNA test | 24,731 | 10,414 | 17.10691 | 0.00581 | 1,791,535 | 830,615 |

Same definitions as Table S1.

**Table S10.** Scenario results and efficiency frontier (LSBE-HGD cohort)

| Strategy | Cost (US$) | Incremental Cost (US$) | Effectiveness (QALY) | Incremental Effectiveness (QALY) | ICER (US$/QALY) | NMB (US$) |
|---|---|---|---|---|---|---|
| Annual AI-assisted endoscopy | 34,829 | Ref | 12.72653 | Ref | Ref | 601,498 |
| AI-assisted endoscopy every 2 years | 35,342 | 513 | 12.52027 | −0.20627 | Absolutely dominated | 590,672 |
| Annual breath test | 35,482 | 653 | 12.77632 | 0.04979 | 13,122 | 603,334 |
| AI-assisted endoscopy every 3 years | 35,511 | 29 | 12.45245 | −0.32387 | Absolutely dominated | 587,112 |
| AI-assisted endoscopy every 4 years | 35,594 | 112 | 12.41923 | −0.35708 | Absolutely dominated | 585,368 |
| AI-assisted endoscopy every 5 years | 35,642 | 160 | 12.39984 | −0.37648 | Absolutely dominated | 584,350 |
| Breath test every 2 years | 35,731 | 249 | 12.54771 | −0.22861 | Absolutely dominated | 591,655 |
| AI-assisted endoscopy every 10 years | 35,732 | 250 | 12.36474 | −0.41158 | Absolutely dominated | 582,505 |
| Breath test every 3 years | 35,813 | 331 | 12.47255 | −0.30377 | Absolutely dominated | 587,814 |
| Breath test every 4 years | 35,853 | 371 | 12.43573 | −0.34059 | Absolutely dominated | 585,934 |
| No surveillance | 35,862 | 380 | 12.26694 | −0.50938 | Absolutely dominated | 577,485 |
| Breath test every 5 years | 35,877 | 395 | 12.41424 | −0.36208 | Absolutely dominated | 584,835 |
| Breath test every 10 years | 35,920 | 438 | 12.37533 | −0.40099 | Absolutely dominated | 582,847 |
| Sponge test every 10 years | 36,326 | 844 | 12.34311 | −0.43321 | Absolutely dominated | 580,830 |
| Sponge test every 5 years | 36,412 | 930 | 12.37045 | −0.40587 | Absolutely dominated | 582,110 |
| Sponge test every 4 years | 36,459 | 977 | 12.38556 | −0.39076 | Absolutely dominated | 582,819 |
| Sponge test every 3 years | 36,539 | 1,057 | 12.41143 | −0.36489 | Absolutely dominated | 584,032 |
| Sponge test every 2 years | 36,702 | 1,220 | 12.46424 | −0.31208 | Absolutely dominated | 586,510 |
| miRNA test every 10 years | 36,848 | 1,366 | 12.38105 | −0.39527 | Absolutely dominated | 582,205 |
| miRNA test every 5 years | 37,058 | 1,576 | 12.42201 | −0.35431 | Absolutely dominated | 584,043 |
| miRNA test every 4 years | 37,172 | 1,690 | 12.44464 | −0.33168 | Absolutely dominated | 585,060 |
| Annual sponge test | 37,197 | 1,714 | 12.62489 | −0.15143 | Absolutely dominated | 594,048 |
| miRNA test every 3 years | 37,367 | 1,885 | 12.48339 | −0.29293 | Absolutely dominated | 586,803 |
| miRNA test every 2 years | 37,762 | 2,280 | 12.56252 | −0.21380 | Absolutely dominated | 590,364 |
| Annual miRNA test | 38,963 | 3,481 | 12.80319 | 0.02687 | 129,566 | 601,196 |

| Strategy | Cost (US$) | Incremental Cost (US$) | Effectiveness (QALY) | Incremental Effectiveness (QALY) | ICER (US$/QALY) | NMB (US$) |
|---|---|---|---|---|---|---|
| Annual AI-assisted endoscopy | 34,829 | Ref | 12.72653 | Ref | Ref | 601,498 |
| Annual breath test | 35,482 | 653 | 12.77632 | 0.04979 | 13,122 | 603,334 |
| Annual miRNA test | 38,963 | 3,481 | 12.80319 | 0.02687 | 129,566 | 601,196 |

Same definitions as Table S1.

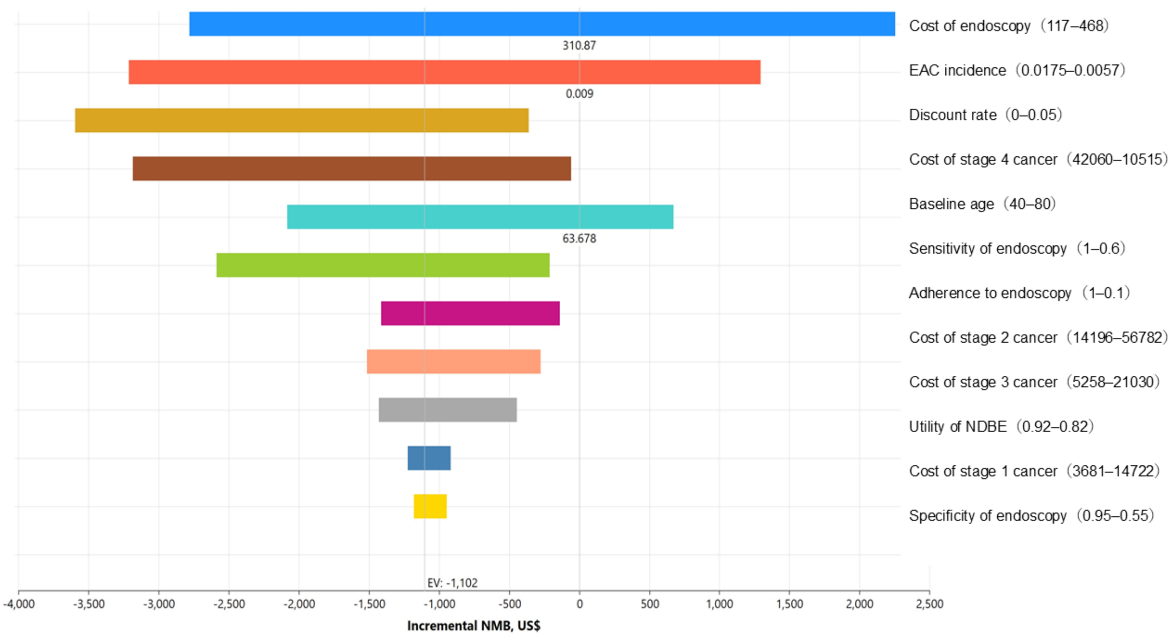


**Figure S1.** Tornado diagram of incremental NMB for LSBE-LGD: No surveillance vs annual endoscopy

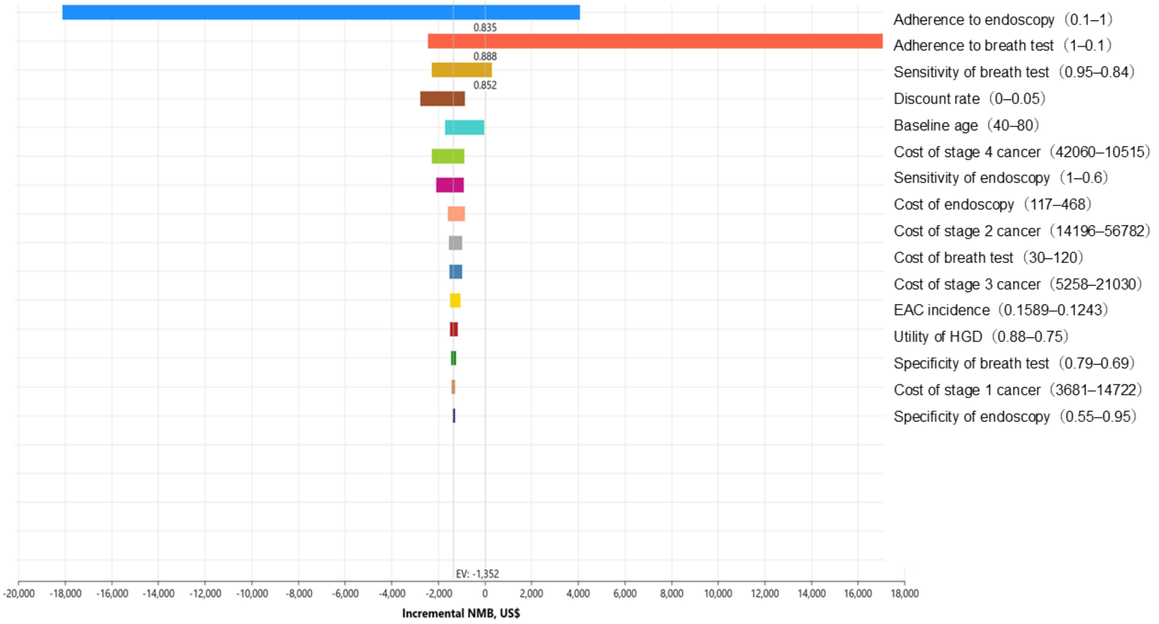


**Figure S2.** Tornado diagram of incremental NMB for LSBE-HGD: Annual endoscopy vs annual breath test

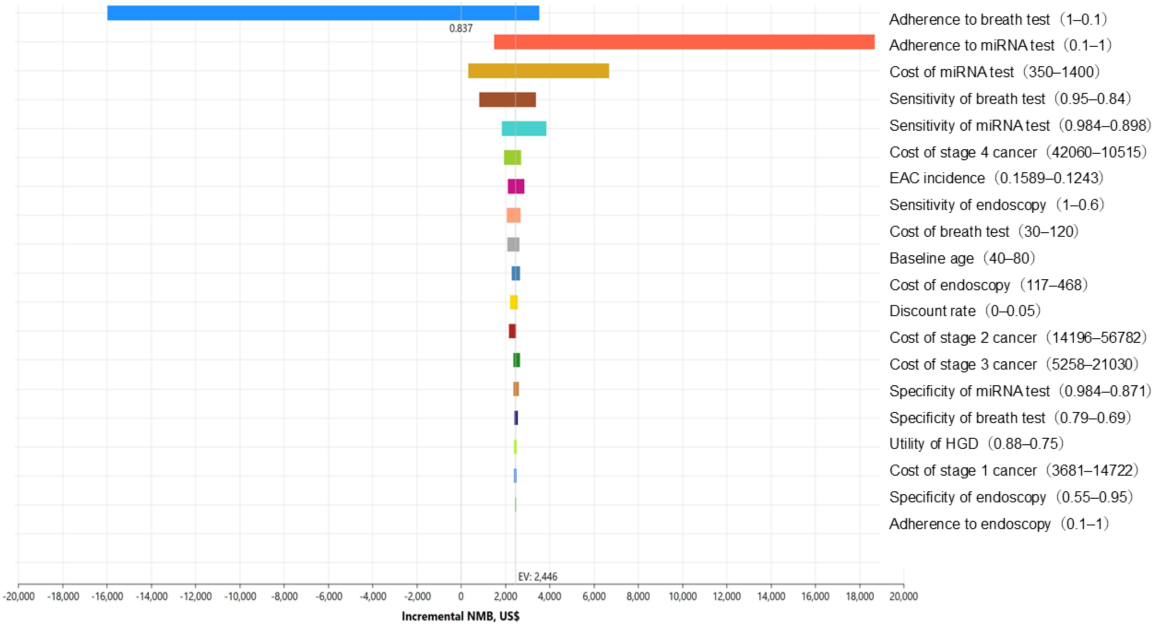


**Figure S3.** Tornado diagram of incremental NMB for LSBE-HGD: Annual breath test vs annual miRNA test

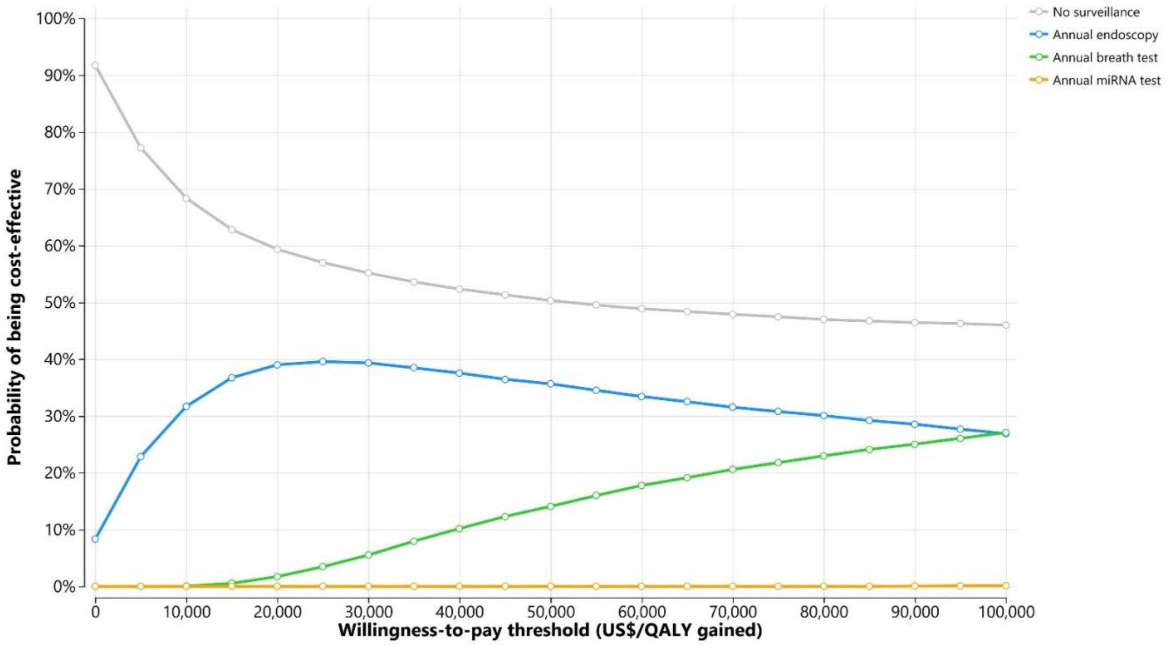


**Figure S4.** Cost-effectiveness acceptability curve for LSBE-LGD including no surveillance

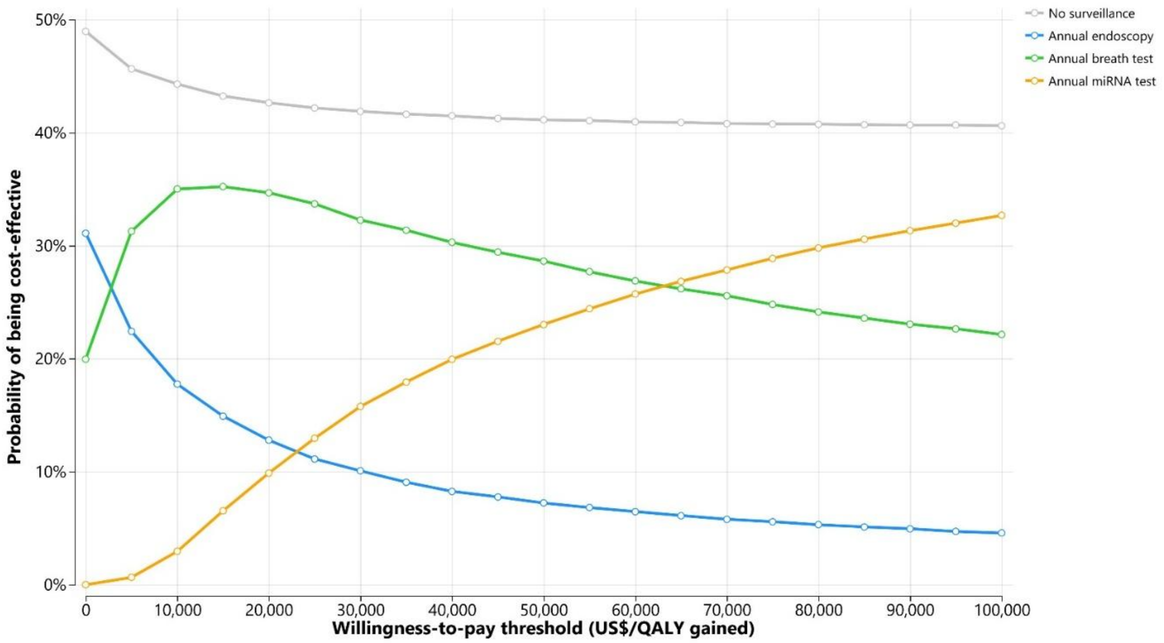


**Figure S5.** Cost-effectiveness acceptability curve for LSBE-HGD including no surveillance

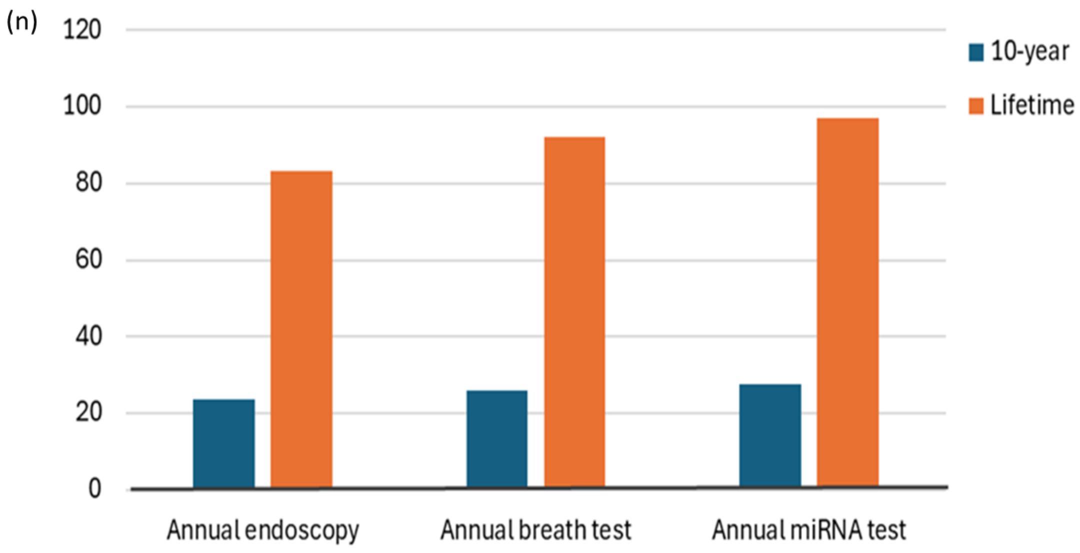


**Figure S6.** EAC deaths averted per 10,000 LSBE-LGD patients over 10 years and over a lifetime

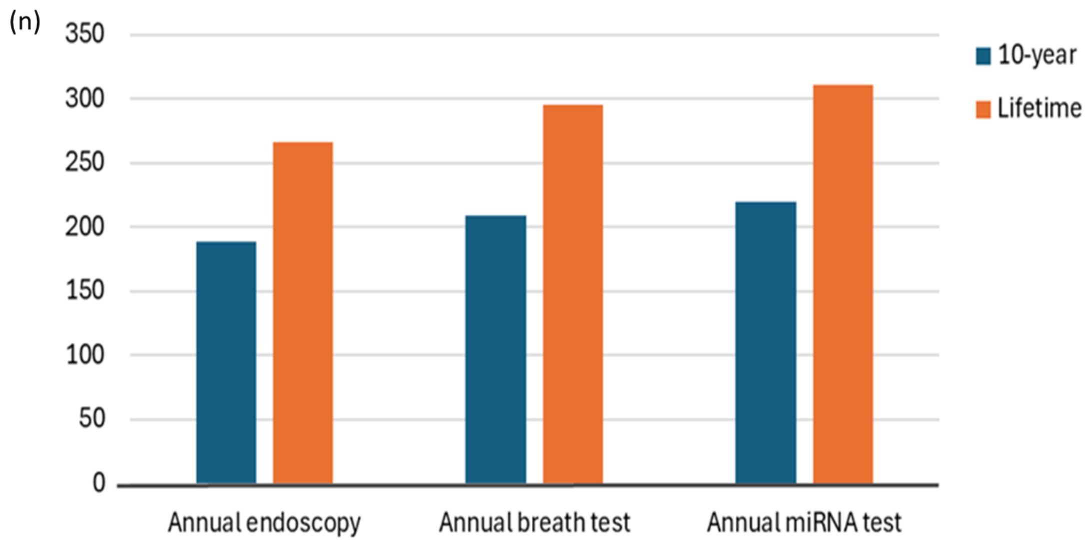


**Figure S7.** EAC deaths averted per 10,000 LSBE-HGD patients over 10 years and over a lifetime

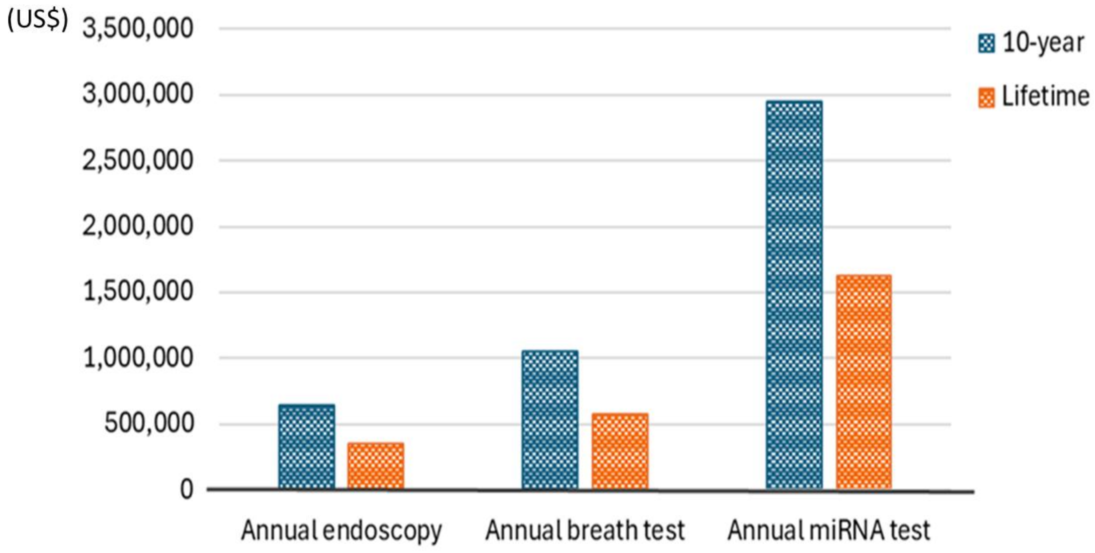


**Figure S8.** Additional costs per EAC death averted among 10,000 LSBE-LGD patients over 10 years and over a lifetime

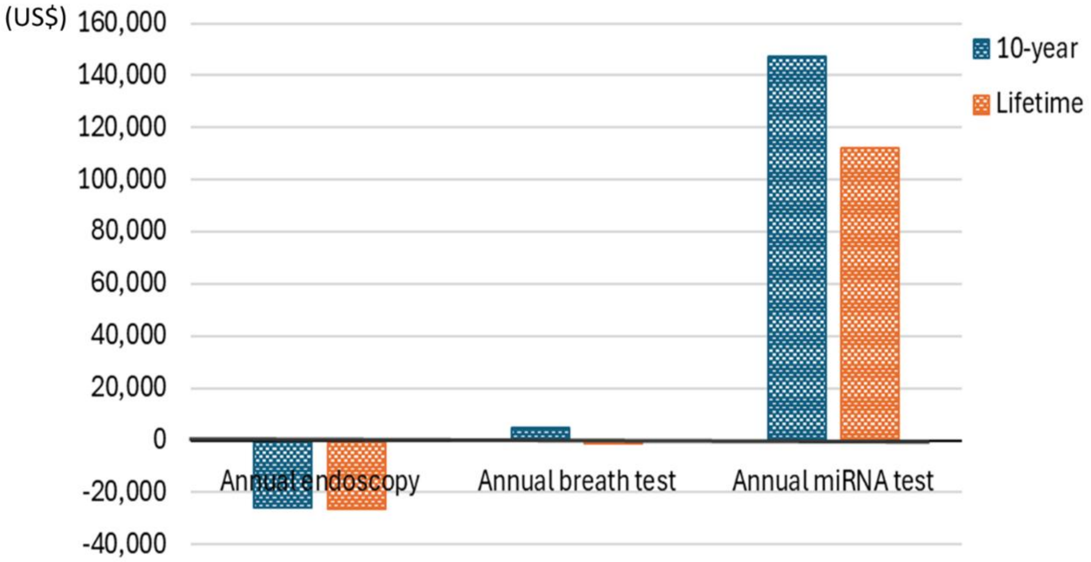


**Figure S9.** Additional costs per EAC death averted among 10,000 LSBE-HGD patients over 10 years and over a lifetime
